# Validating atlas-based lesion disconnectomics in multiple sclerosis: a retrospective multi-centric study

**DOI:** 10.1101/2021.05.03.21256161

**Authors:** Veronica Ravano, Michaela Andelova, Mário João Fartaria, Mazen Fouad A-Wali Mahdi, Bénédicte Maréchal, Reto Meuli, Tomas Uher, Jan Krasensky, Manuela Vaneckova, Dana Horakova, Tobias Kober, Jonas Richiardi

## Abstract

The translational potential of MR-based connectivity modelling is limited by the need for advanced diffusion imaging, which is not part of clinical protocols for many diseases. In addition, where diffusion data is available, brain connectivity analyses rely on tractography algorithms which imply two major limitations. First, tracking algorithms are known to be sensitive to the presence of white matter lesions and therefore leading to interpretation pitfalls and poor inter-subject comparability in clinical applications such as multiple sclerosis. Second, tractography quality is highly dependent on the acquisition parameters of diffusion sequences, leading to a trade-off between acquisition time and tractography precision.

Here, we propose an atlas-based approach to study the interplay between structural disconnectivity and lesions without requiring individual diffusion imaging. In a multi-centric setting involving three distinct multiple sclerosis datasets (containing both 1.5T and 3T data), we compare our atlas-based structural disconnectome computation pipeline to disconnectomes extracted from individual tractography and explore its clinical utility for reducing the gap between radiological findings and clinical symptoms in multiple sclerosis. Results using topological graph properties showed that overall, our atlas-based disconnectomes were suitable approximations of individual disconnectomes from diffusion imaging. Small-worldness was found to decrease for larger total lesion volumes thereby suggesting a loss of efficiency in brain connectivity of MS patients. Finally, the global efficiency of the created brain graph, combined with total lesion volume, allowed to stratify patients into subgroups with different clinical scores in all three cohorts.

## 1. Introduction

The increased availability and quality of non-invasive brain connectivity imaging, using either diffusion or functional MRI data, coupled with methodological advances, has triggered the emergence of subfields at the intersection of neuroscience and network science. In network neuroscience (Bassett and Sporns, 2017), graphs are used to model connections between brain regions, enabling an arsenal of mathematical methods which facilitate systems-level thinking and thus new ways of investigating the organization of the brain (Bullmore and Bassett, 2011; Richiardi et al., 2013; Sporns et al., 2005). Compared to diffusion MR images or tractograms, using more interpretable high-level representations of structural connectivity, such as brain graphs and their underlying topological features, enables more reliable inter-subject comparisons (Meskaldji et al., 2013).

During the last decade, the study of structural connectomes has provided a better and more extensive understanding of several neurological and psychiatric diseases. The ‘disconnectome’ approach combines the study of lesion location with structural connectomics to investigate the impact of resulting disconnections (Catani et al., 2012; Foulon et al., 2018; Fox, 2018). Where disruption of brain connectivity is found to affect higher functions, the condition can be described as a “disconnection syndrome” (Carrera and Tononi, 2014; Catani and Ffytche, 2005).

Recently, repeated findings have led multiple sclerosis (MS) to be also characterized by structural disconnectivity (Rocca et al., 2015), on top of the well-known main drivers of the disease (i.e. grey matter pathology, brain atrophy, diffuse microstructural damage of normal appearing white matter). Brain damage in MS involves the transection of white matter tracts by lesions, creating a disconnection syndrome which is associated with a specific clinical phenotype (Llufriu et al., 2017). T2 hyperintense lesions might lead to remote axonal damage by both antero- and retrograde degeneration (“dying back“ and Wallerian degeneration (Dziedzic et al., 2010; Lucchinetti et al., 2000)). Axonal loss has been identified as the major determinant of irreversible neurological disability in MS patients (Hayes and Ntambi, 2020). An accelerated rate of brain atrophy in MS patients, together with disruption of nerve signals in the central nervous system, can cause multiple symptoms that vary widely from one patient to another. In particular, connectivity studies have shown that the overall efficiency of functional connectomes is affected by the disease (Yaou et al., 2017) (Rocca et al., 2016)(Fleischer et al., 2019) and that small-worldness (defined as the ratio between clustering coefficient and the characteristic shortest path length) seems to dissipate with increasing lesion load (Faivre et al., 2016). Further, a decrease in graph efficiency was shown to correlate with EDSS, disease duration and total WM lesion loads (Shu et al., 2011) therefore showing the potential of such new quantitative features to help narrowing the clinico-radiological paradox (Barkhof, 2002).

Although tractography has been proven to be in good accordance with ex-vivo histological experiments (Buckner et al., 2011; Donahue et al., 2016), its use in clinical settings suffers from several pitfalls. Notably, the diffusion signal strength and the derived tracking algorithm outcome are highly dependent not only on physiological factors and on the way axons lay in a given voxel, but also on the image reconstruction methods and acquisition parameters chosen. The quantitative interpretability of the results is therefore highly dependent on model assumptions (Jones et al., 2013) and renders inter-subject comparisons more difficult and less reliable. Moreover, tracking algorithms are highly sensitive to two major time-consuming acquisition variables: spatial resolution and q-space coverage (Calabrese et al., 2014). As acquisition time is a critical limitation in clinical protocols, diffusion imaging sequences fulfilling these high standards are rarely acquired. Other technical limitations of tractography include the impact of white matter lesions on diffusion properties (Tievsky et al., 1999) that can in turn interfere with the tracking algorithm (Ciccarelli et al., 2008), although steady progress is being made to mitigate these detrimental effects (Lin et al., 2005; Lipp et al., 2020; Pawlitzki et al., 2017; Reich et al., 2007).

To reduce inter-subject variability and provide higher quality estimate of structural connectivity compared to classical clinical settings, previous studies proposed to model individual structural brain disconnectivity without requiring diffusion imaging nor tractography (Griffis et al., 2021; Ravano et al., 2020, 2019). Structural connectivity is approximated using the HCP842 atlas (Yeh et al., 2018), a population-averaged tractography atlas built using 842 healthy controls of the Human Connectome Project (Van Essen et al., 2012). Of interest, previous studies showed the validity of using a connectome atlas to approximate normal population (Horn et al., 2016). Similarly to what was proposed by (Griffis et al., 2021), we estimate individual connectivity loss resulting from white matter lesions by overlapping an automatically generated lesion mask with the tractography atlas and using brain graphs to model an individual structural disconnectome: a brain map representing the affected neuronal connections. In this retrospective multi-centric study, we evaluate the proposed approach, by comparing it as closely as possible with disconnectomes extracted from individual diffusion-based tractography in a subset of patients and we explore its clinical usefulness in reducing the clinico-radiological paradox in MS.

## 2. Datasets

Demographics and relevant MR protocol parameters are reported in Table 1 for all cohorts. The number of patients in each dataset corresponds to those fulfilling the applicability criterion defined in Section 3.2.3.

**Table 1.** Demographics and relevant MR protocols for the three datasets. Lesion volumes are estimated from manual segmentations when available (Diffusion and 1.5T cohorts) and from LeMan-PV otherwise (3T cohort).

|  |  | Diffusion cohort | 1.5T cohort | 3T cohort |
| --- | --- | --- | --- | --- |
| **N (Males)** |  | 41 (13) | 208 (69) | 496 (158) |
| **Age [years]** |  | 35.1 ± 9.9 | 28.3 ± 7.3 | 28.0 ± 8.2 |
| **Median EDSS [min,max]** |  | 1.5 [1, 4] | 1.5 [0, 3.5] | 2 [0, 7.5] |
| **Disease Duration [months]** |  | 24 ± 18 | < 3 | 143 ± 79 |
| **Lesion Load [mL]** |  | 7.7 ± 9.4 | 4.2 ± 5.3 | 13.0 ± 14.6 |
| **MR System** | Vendor | Siemens | Philips | Siemens |
|  | Scanner | 3T Trio | 1.5T Gyroscan | 3T Skyra |
| **T1w** | Voxel size [mm<sup>3</sup>] | 1x1x1.2 | 1x1x1 | 1x1x1 |
|  | Acquisition Type | 3D | 3D | 3D |
|  | TR/TE/TI [ms] | 2300/2.98/900 | 25/5/- | 2300/2.96/900 |
|  | Flip Angle | 9° | 30° | 9° |
| **FLAIR** | Voxel size [mm<sup>3</sup>] | 1x1x1.2 | 1x1x1.5 | 1x1x1 |
|  | Acquisition Type | 3D | 2D | 3D |
|  | TR/TE/TI [ms] | 5000/394/1800 | 11000/140/2600 | 5000/397/1800 |
|  | Flip Angle | 120° | 90° | 120° |
| **DSI** | Voxel size [mm<sup>3</sup>] | 1x1x3 | - | - |
|  | TR/TE [ms] | 8600/144 | - | - |
|  | b-max [s/mm<sup>2</sup>] | 8000 | - | - |

### 2.1. Diffusion Cohort: controls and patients with diffusion data

Forty-five patients with relapsing-remitting MS (disease duration < 5 years) were scanned on a 3T MRI system (MAGNETOM Trio, Siemens Healthcare, Erlangen, Germany) using a 32-channel head coil at the Lausanne University Hospital (Simioni et al., 2014). The acquisition protocol included: (i) high-resolution magnetization prepared - rapid gradient echo (MPRAGE), (ii) fluid attenuation inversion recovery (FLAIR) and (iii) diffusion spectrum imaging (DSI).

T2 hyperintense lesions on FLAIR were manually marked by one radiologist and one neurologist, and a consensus lesion mask was created as described in (Fartaria et al., 2016).

Two patients were discarded due to incomplete imaging data and two other patients did not fulfil the applicability criterion resulting in a subset of forty-one MS patients (13 males, average age 35.1+/-9.86, EDSS ϵ [1,4], median EDSS 1.5) (see Section 3.2.3 for further details). All patients provided written informed consent, and the study was approved by the ethics committee of the State of Vaud, Switzerland.

### 2.2. 1.5T cohort

An observational Study of Early Interferon beta 1-a Treatment in high risk subjects after clinical isolated syndrome (SET study (Horakova et al., 2013)) recruited 220 patients within four months after their first clinical event suggestive of MS and all eventually converted to clinically definite MS. Patients were scanned at 1.5T (Gyroscan NT 15, Philips Healthcare, Best, The Netherlands) and clinically evaluated every six months during a period of four years. The study involved eight centers and was coordinated by the Charles University Hospital of Prague, where all patients underwent MR examinations. The acquisition protocol included i) FLAIR and ii) T1-weighted spoiled-gradient recalled (SPGR).

An expert neurologist with nine years of experience manually segmented T2 hyperintense lesions in FLAIR. A subset of 208 patients was retained for analysis following our applicability criterion (69 males, average age 28+/-7.3, EDSS ϵ [0,3.5], median EDSS 1.5) (see Section 3.2.3).

### 2.3. 3T cohort

Five hundred eighty-nine patients affected by any form and stage of MS were recruited in a project (the Spinal Cord Grant (SCG)) coordinated by the Charles University Hospital of Prague (Czech Republic). Patients underwent yearly clinical examination and MR imaging at 3T (MAGNETOM Skyra, Siemens Healthcare, Erlangen, Germany). The acquisition protocol included high-resolution MPRAGE and 3D FLAIR.

Due to the considerable number of patients, manual segmentations of white matter lesions were not available, but automated lesion segmentation was performed as detailed in Section 3.1.

Four hundred ninety-six patient datasets fulfilled the applicability criterion (158 males, average age 28 +/- 8.2, EDSS ϵ [0, 7,5], median EDSS 2). Among those, 74.4% were affected by the relapse-remitting form of the disease, 14.6% had clinically isolated syndrome, 10.3% secondary progressive, 0.4% primary progressive and 0.2% progressive-relapsing.

One hundred thirty-two patients were present in both the 1.5T and the 3T cohorts, but were scanned at different time points (on average 10.8 ± 1.08 years apart, always examined on the 1.5T scanner first). On average, patients experienced an EDSS increase of 0.28 ± 1.2 between the two scanning sessions. However, the EDSS difference varied greatly among the cohort, ranging between -2.5 and 4.0.

All participants in the 1.5T and the 3T cohorts agreed with collecting and retrospectively analyzing their clinical, immunological and MRI data within an international, online registry and platform for collecting prospective data on patients with MS (MSBase) (Butzkueven et al., 2006), and within the Czech national registry of MS patients (ReMuS). Therefore, neither ethics committee approval nor separate informed consent were obtained for this study.

## 3. Methods

Due to patient privacy, the clinical data used in this study cannot be made openly available. The Python code used to extract the disconnectome graph and relevant topological metrics from a lesion mask normalized to MNI space is publicly available (https://gitlab.com/acit-lausanne/lesion-disconnectomics), together with pointers to relevant open data as examples. An R based visualisation script is also made available.

### 3.1. Pre-processing

Lesions were segmented using the LeMan-PV prototype, an automated lesion segmentation technique that performs segmentations on FLAIR and T1-weighted contrasts in two main steps: lesions are first detected using a supervised voxel-wise approach (Fartaria et al., 2016) and then delineated based on partial volume estimation (Fartaria et al., 2017). Lesion concentration maps resulting from LeMan-PV were binarized following the methodology described in (Fartaria et al., 2017). LeMan-PV segmentations were computed for the diffusion cohort (in addition to manual segmentation) and the 3T cohort, whereas it was not applied to the 1.5T cohort as the data were not compliant with the minimal requirements defined by LeMan-PV for a good segmentation quality (in particular, data were acquired using a 2D rather than a 3D FLAIR sequence)(Fartaria et al., 2016).

To overlay the lesion mask with the tractography atlas, a non-rigid spatial registration was used to transform the native T1-weighted contrast of each patient to the standard T1-weighted MNI152 (2009a) template space using b-spline trilinear interpolation from the Elastix v.4.800 implementation (Klein et al., 2010) (the parameter files are available in the code repository). The transformation was then applied to the binary lesion mask with nearest neighbour interpolation to ensure binary output mask and preserving lesion topology.

Patients’ normalised brains were then segmented in 274 fine-grained subregions using the Brainnetome parcellation atlas (Fan et al., 2016), a connectivity-based atlas extracted from multimodal neuroimaging data from 40 healthy subjects.

### 3.2. Modelling structural disconnectomes from a tractography atlas

Similarly to the method described in (Griffis et al., 2021), we estimate disconnectomes without diffusion imaging data, using a population-averaged structural tractography atlas, namely the HCP842 tractography atlas (Yeh et al., 2018). The HCP842 atlas was built by averaging the Spin Distribution Function (SDF) in each voxel for 842 healthy subjects from the Human Connectome Project (Van Essen et al., 2012) (372 males, age range between 22 and 36 years old) whose diffusion data were reconstructed in MNI space using a q-space diffeomorphic strategy (Yeh and Tseng, 2011).

#### 3.2.1. Computation of the individual disconnectomes

For each patient, affected connections were extracted by isolating the atlas streamlines passing through the patient’s normalised lesion mask using DSI Studio (http://dsi-studio.labsolver.org). Subsequently, the connectivity of affected connections between all possible pairs of brain regions (*i, j*) extracted from the Brainnetome atlas was modelled as an adjacency matrix of damaged connectivity *A*_*c*_. Each element of the matrix *A*_*C*_(*i, j*) represented the number of damaged streamlines passing through the corresponding pair of regions (*i, j*). The reference “healthy” brain connectivity was represented as an adjacency matrix *A*_*t*_ where elements *A*_*t*_(*i, j*) represented the number of streamlines passing through the corresponding pair of regions (*i, j*) in the tractography atlas (see Figure 1A to C and Figure2A to B). Note that *A*_*t*_ is the same for all patients, while *A*_*c*_ is patient-specific and depends on lesions.

**Figure 1.**
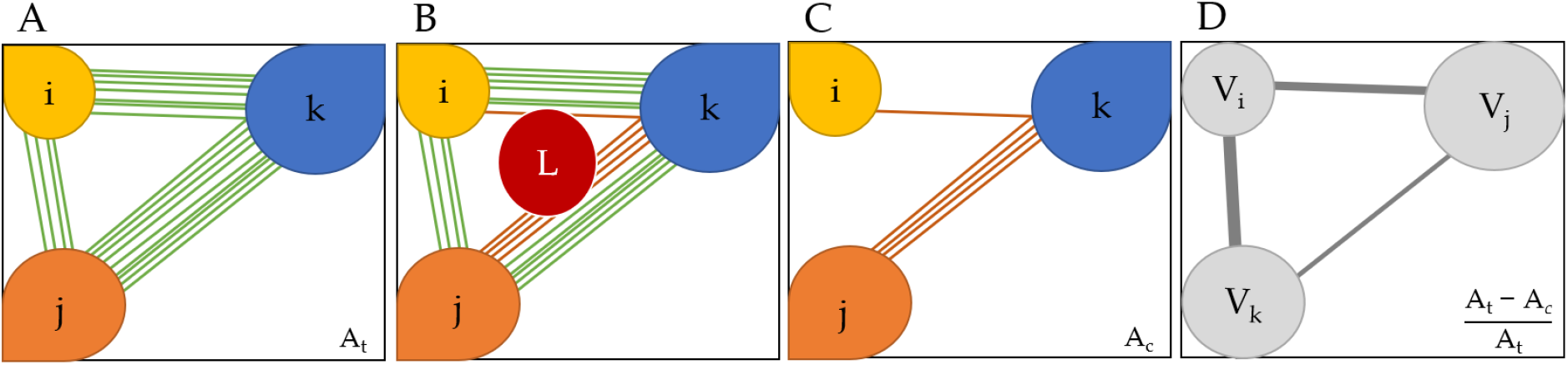
Extraction and modelling of a disconnectome. **A.** Simplified representation of the atlas-based tractogram connectivity of three brain regions i, j and k and their respective streamlines. At is the associated adjacency matrix of the atlas connectome, where each element A_t_(i,j) represents the number of streamlines connecting the pair of regions (i,j). **B.** Overlapping of the atlas tractogram with the lesion mask. Streamlines passing through the lesion L are highlighted in red. **C.** Affected streamlines are isolated. A_c_ is the adjacency matrix of the affected streamlines, where each element A_c_(I,j) represents the number of affected streamlines connecting the pair of regions (i,j) **D.** The brain graph representation where brain areas i, j and k are represented respectively by nodes V_i_ V_j_ and V_k_ and edges are weighted by the relative number of affected streamlines. The adjacency matrix of the remaining connectivity is defined as the relative difference between A_t_ and A_c_.

**Figure 1.**
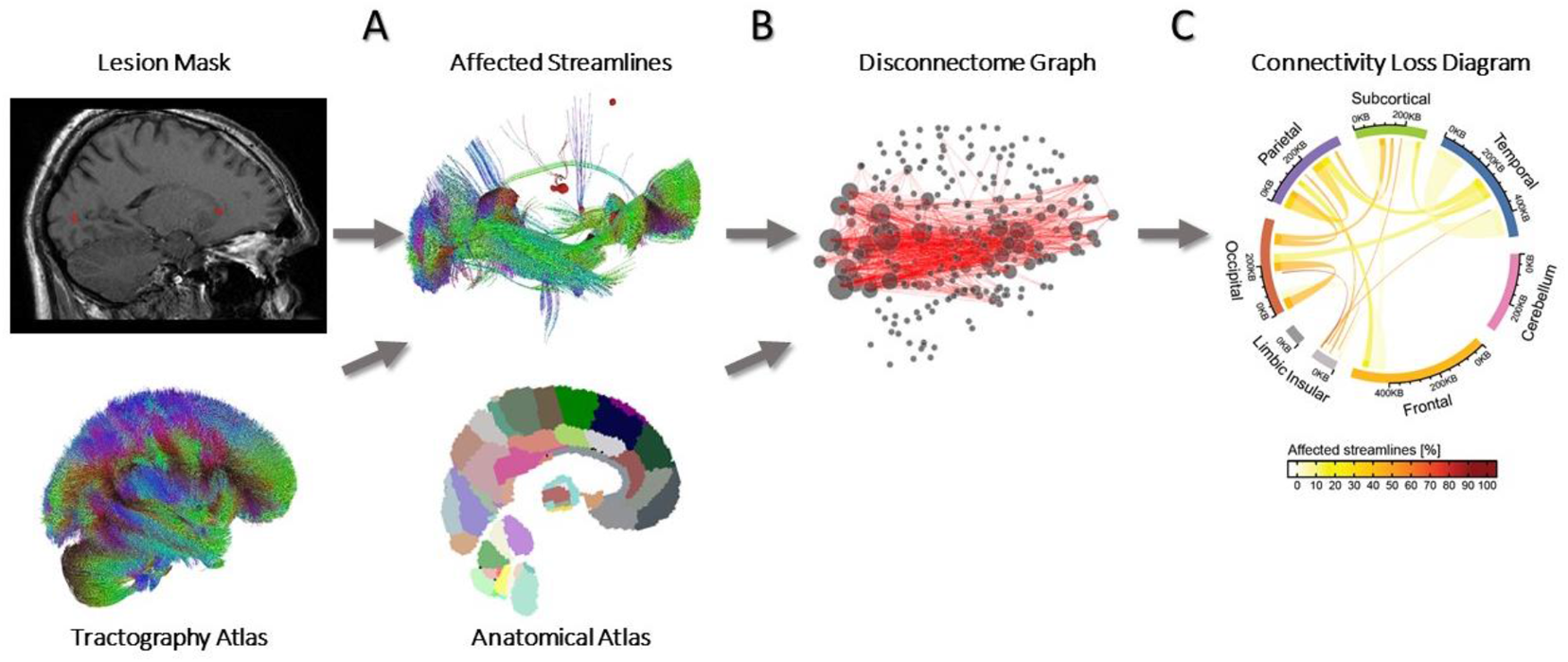
Extraction and representation of a disconnectome. A. The streamlines intersecting the lesion mask are isolated from the healthy tractography atlas. B. Affected streamlines are overlaid with an anatomical atlas to create a brain graph representation of disconnectivity. C. Lobe-wise summary representation of disconnectivity, visualized using Circos (Connors et al., 2009).

**Figure 2.**
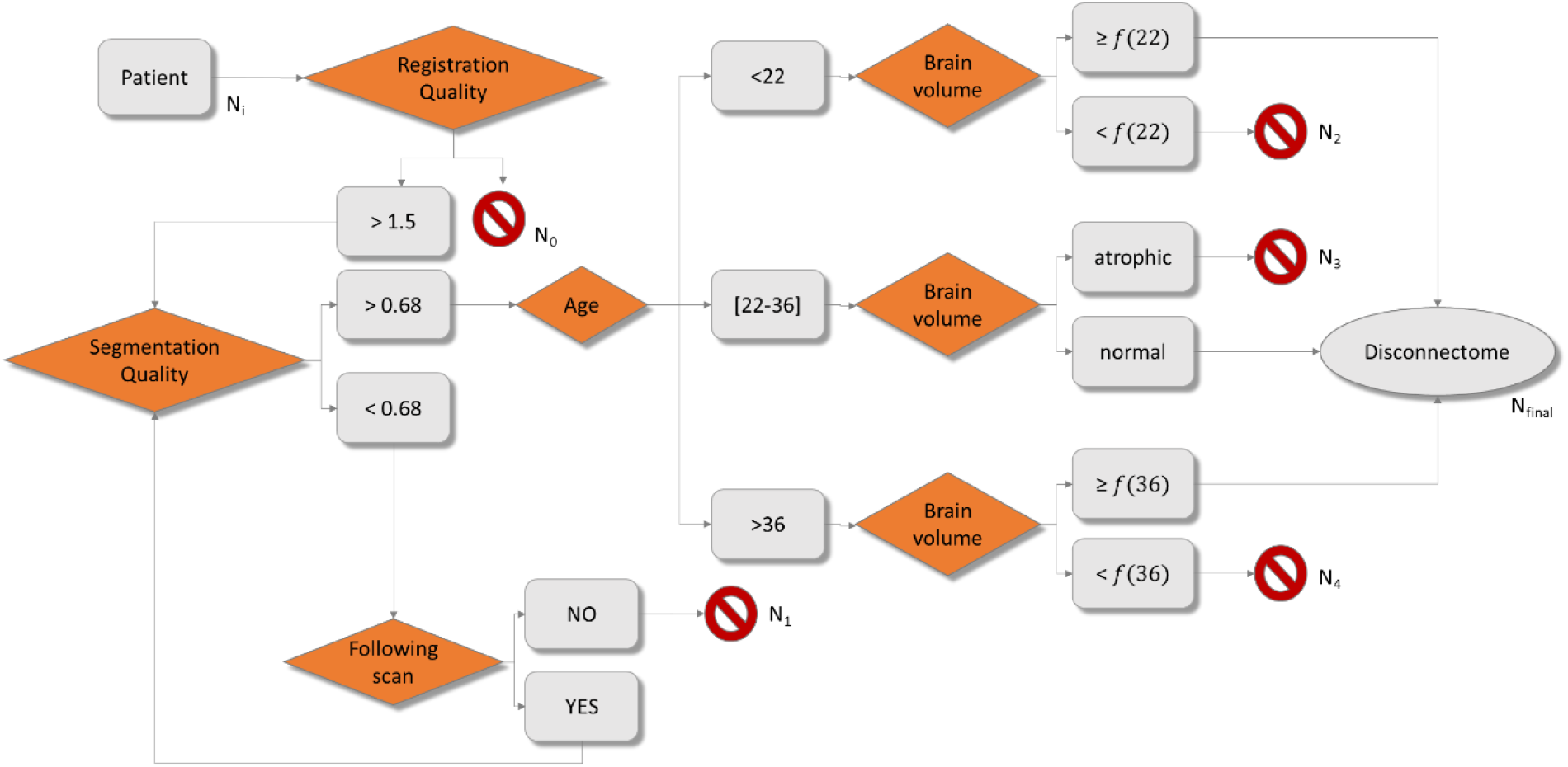
Flow diagram showing the applicability conditions of the atlas-based disconnectome model, based on age, brain atrophy and brain segmentation quality estimated from MorphoBox (Schmitter et al., 2015). f(x) is the 3% percentile of normative ranges. Ni: initial number of patients. N0: number of patients discarded due to poor registration quality. N1: number of patients discarded due to bad segmentation quality. N2: number of patients younger than the age range covered by the tractography atlas, who are discarded due to abnormally low brain volume. N3: number of patients inside the atlas age range discarded due to abnormally low brain volume. N4: number of patients older than the age range covered by the tractography atlas, who are discarded due to abnormally low brain volume.

The matrix of affected connectivity *A*_*c*_ was then used to build a brain graph where each parcellation area was represented by one node *V*_*i*_. Each edge *E*(*i, j*) represented the percentage of streamlines affected by lesions connecting areas i and j with respect to the atlas connectivity *A*_*t*_(*i, j*):

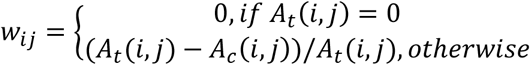

This weighted graph model ensures the number of nodes is consistent across patients and that the edge weights lie between 0 and 1 (see Figure 1D), where higher percentages of affected streamlines result in lower weight values.

For visualization, a simplified diagram of connection loss was obtained by grouping Brainnetome regions into the main brain lobes and displaying them using Circos (Connors et al., 2009): the line thickness represents the number of affected streamlines whereas the colour codes for the percentage of affected streamlines relative to the healthy atlas (see Figure 2C).

#### 3.2.2. Extraction of Graph Features

One way of using graphs for statistical analysis or machine learning is to represent them as vectors, i.e., to use graph embedding. Then, a vast number of algorithms, both classical and deep, can be used without modification for predictive modelling on graphs (Richiardi et al., 2013). Embedding techniques include the extraction of topological features characterizing graph properties.

Topological graph features can be defined at different scales: at large-scale, they reflect properties on the entire graph (e.g. small-worldness), at intermediate scale, the metrics are computed on subgraphs (e.g. edge betweenness) and at small scale they reflect node features (e.g. node strength) (De Vico Fallani et al., 2014). Here, we extracted different small- and large-scale features computed with the NetworkX Python library (Hagberg et al., 2008) to study both the performance of our atlas-based method compared to individual disconnectomes, and the usefulness of these metrics for clinical applications. The topological features used are described in Table 2.

**Table 2.**
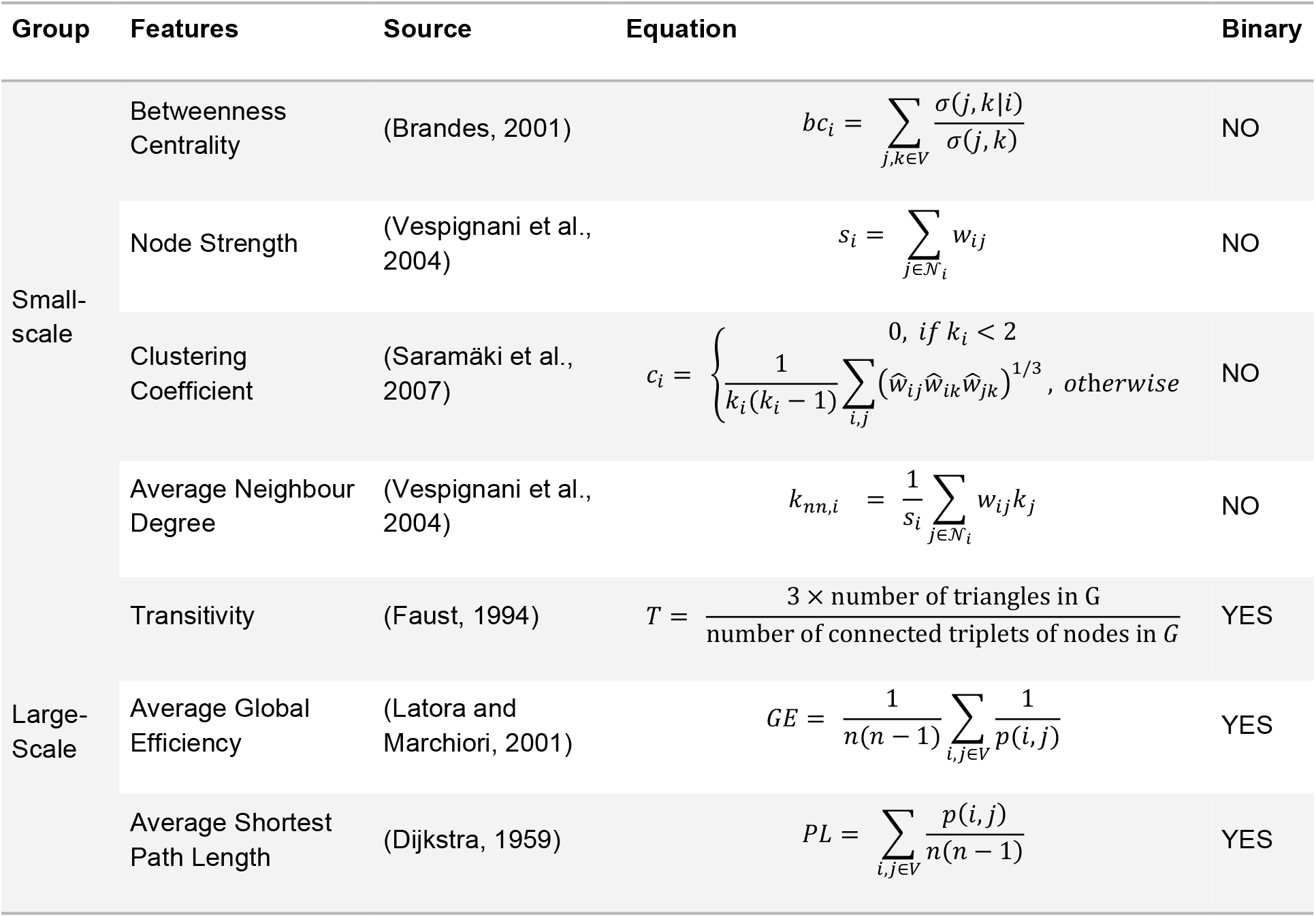
Description of topological metrics extracted from the disconnectome graph. In the following equations *V* is the set of nodes, *σ*(*j, k*) is the number of shortest paths between nodes *j* and *k, σ*(*j, k*|*i*) is the number of those paths passing through node *i*, 𝒩_*i*_ is the set of nodes in the neighbourhood of node *i*,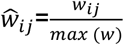 and *k*_*i*_ the node degree defined as the number of edges connecting to node *i. p*(*i, j*) the shortest path length between nodes *i* and *j* and with n=274 the number of nodes in the graph.

In terms of large-scale features, we use transitivity (T), global efficiency (GE), and path length (PL) as defined for binary graphs, and the disconnectome graphs were binarized using a threshold τ on edge weights so that the most severely affected connections were modelled as totally disrupted, and the mildly affected ones were modelled as intact. Binary edges were therefore defined such as:

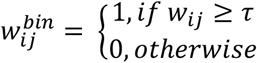

We checked that graphs remained connected after thresholding and artificially added one edge to reconnect eventual disconnected components. A detailed analysis aiming at defining the optimal binarization threshold for disconnectome graphs is described in Supplementary Materials Appendix B and resulted in a threshold of 0.7.

Further, we extract small-scale features for each node i, such as its strength (*s*_*i*_), its neighbour degree (*k*_*nn,i*_), clustering coefficient (*c*_*i*_) and betweenness centrality (*bc*_*i*_). These metrics were also averaged across all nodes to provide additional insights on large-scale topology.

#### 3.2.3. Model Assumptions and Applicability Conditions

Our proposed model relies on the assumption that a patient’s brain connectivity can be estimated from the HCP842 atlas. However, the effects of brain atrophy on axonal projections are unclear and might lead to interpretation issues when approximating atrophic brains with the HCP842 atlas. In fact, brain atrophy, resulting from the loss of neurons or neuronal connections, potentially leads to substantial quantitative changes in structural connectivity (Kuceyeski et al., 2015).

To be coherent with our initial assumption and avoid possible misinterpretation resulting from the presence of atrophy, we used an applicability criterion to discard unsuitable patients. As brain atrophy can be caused either by normal aging or pathology, we implemented a criterion based on both age and brain volume relative to total intracranial volume (TIV) estimated with the MorphoBox prototype brain morphometry software (Schmitter et al., 2015). Further, our method heavily relies on spatial registration of images in MNI space. Therefore, to assess the accuracy of the spatial alignment, we implemented a registration quality check based on the metric optimized by the Elastix algorithm, namely the mutual information. The quality of aligned images associated with the lowest scores was visually assessed in each cohort, and we concluded all registrations were satisfactory. Based on this observation, a mutual information threshold was set to 1.5.

The quality of the MorphoBox segmentation was assessed to discard patients with unreliable brain volume estimates. The segmentation quality was computed as the correlation between the grey matter a posteriori (GM) probability map generated by MorphoBox and a priori GM probability map. Correlations higher than 0.68 were thought to reflect high segmentation quality according to (Chow and Paramesran, 2016). In case of low segmentation quality, brain volume and segmentation quality were assessed for scans in subsequent timepoints and the first scan with sufficiently high segmentation quality was retained as reference. Then, age-and gender-matched MorphoBox reference ranges were used to detect abnormal brain volume in patients belonging to the tractography atlas age range (22 to 36 years old). The 3% percentile *f* (*x*) of normative ranges was chosen not to be too conservative as the sharp boundaries of the atlas age range were already considered. Thresholds were defined for patients outside this age range. In particular, patients below twenty-two and above thirty-six years old were identified as suitable if their brain volume was above *f* (22) and *f* (36), respectively.

A flow diagram showing the applicability criterion of our proposed method is represented in Figure 3. The precise breakdown of datasets after applying our applicability conditions is reported in Supplementary Table 1.

**Figure 3.**
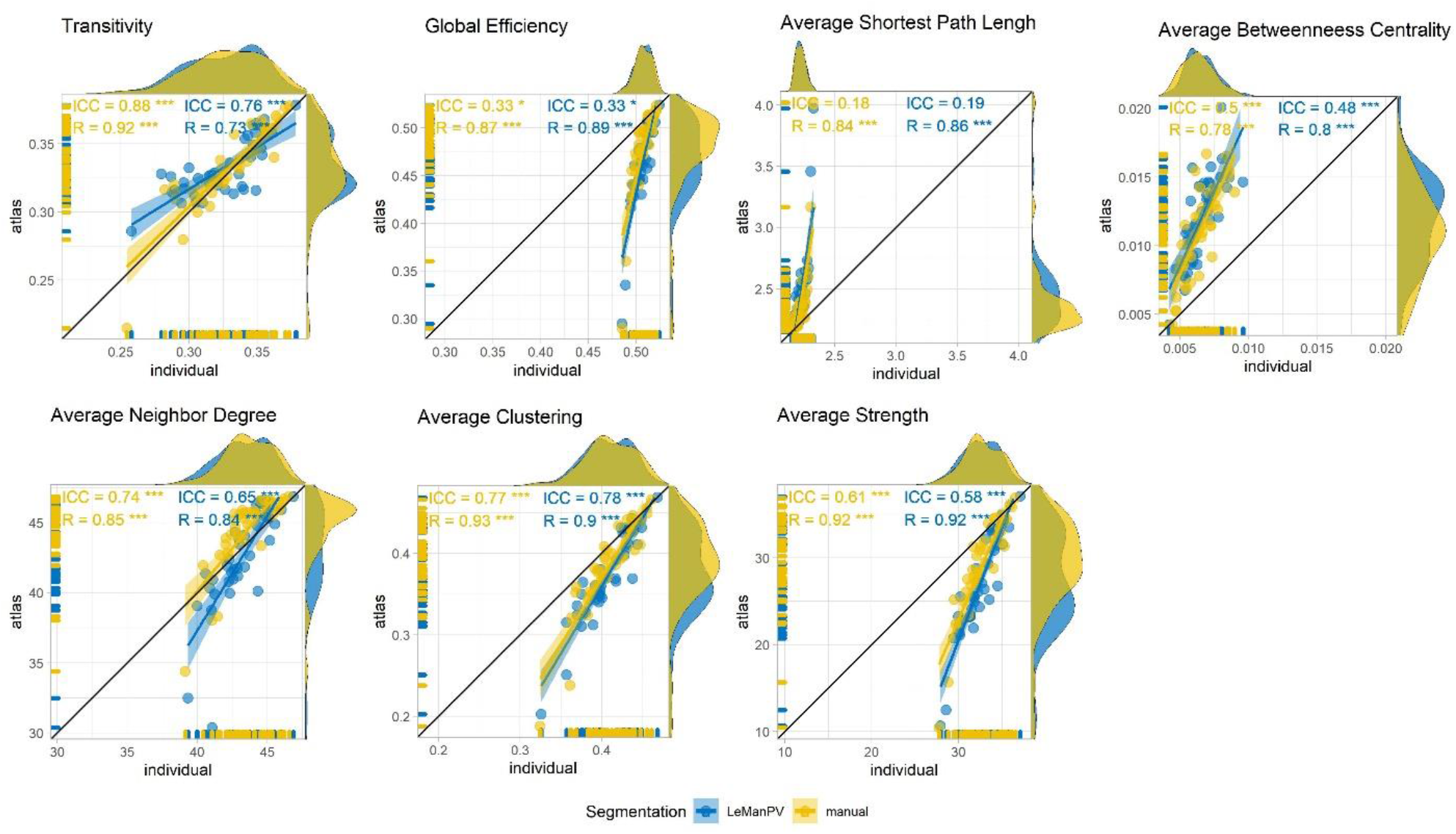
Large scale topological features extracted from atlas-based disconnectome graph (y-axis) plotted against equivalent metrics extracted from individual tractography-based disconnectome (x-axis) for patients of the diffusion cohort. Each point is a patient. Disconnectome features derived from manual lesion segmentation are shown in yellow, and automated lesion segmentation with LeManPV in blue. ICC(3,1) and Spearman’s correlation are reported with their respective significance level after correction for multiple comparison. * p<0.05, ** p<0.01, *** p<0.001

### 3.3. Comparison with disconnectomes based on individual tractography

Individual variability in brain connectivity in patients can be either caused by individual anatomical factors or by pathological phenomena such as brain atrophy, oedema, or white and gray matter lesions, degree of de- and remyelination. To estimate the error made under the assumption that patients’ brain connectivity could be approximated with the population-averaged HCP842 tractography atlas, we compared atlas-based disconnectome graphs to the equivalent graphs derived from individual tractography in our diffusion cohort.

To build individual whole-brain tractograms, the same reconstruction strategy and tracking algorithm were used as described in (Yeh et al., 2018), in order to ensure consistency with the HCP842 tractography atlas. All reconstruction and tracking parameters were chosen following (Yeh et al., 2018) and are described in Supplementary Materials Appendix: A. The disconnectome was then extracted and modelled following the same procedure as shown in Section 3.2.1 and will be hereafter referred to as individual tractography-based disconnectome, as opposed to the atlas-based disconnectome.

A connection-level comparison of disconnectomes was first performed. To this end, lobe-wise connections in the atlas-based and individual tractography-based disconnectome matrices were ranked according to their connectivity strength in the diffusion cohort. Then, rank-rank Spearman correlation was computed between the two disconnectome models for both lesion segmentation techniques.

Correlations between large-scale topological metrics extracted from both disconnectome models were investigated on the population level in the diffusion cohort by computing the intraclass correlation coefficient (ICC(3,1)) and Spearman’s correlation. The ICC reflects the agreement between the two methods, whilst Spearman correlation is insensitive to systematic over or underestimations but indicates to what extent the ranks of the data are preserved.

Small-scale topological features were extracted for all 274 nodes from atlas-based and individual tractography-based disconnectomes and the intraclass correlation ICC(3,1) and Spearman correlation were computed for each patient in the diffusion cohort and each lesion segmentation strategy. The impact of total lesion volume on patient-level correlations was also studied.

Finally, to investigate the possible impact of patients’ age, brain atrophy and segmentation quality on our validation analysis, Spearman correlations were computed between these metrics and the relative absolute topological differences (between atlas-based and individual tractography-based disconnectomes).

### 3.4. Clinical Applications in Multiple Sclerosis

To test the clinical relevance of our technique, we investigated the potential of atlas-based disconnectome features to reduce the gap between radiological findings and physical disability in MS.

First, to verify that the extracted topological features were related to actual physiological phenomena instead of measurement noise, the variation of large-scale topological features with total lesion volume was analysed in all cohorts. The analysis was restricted to patients with a TLV smaller than 50 mL to provide a comparable range of TLV between datasets and therefore discarding 15 patients from the 3T cohort. Then, the univariate Spearman’s correlation between these features and EDSS was investigated cross-sectionally and compared to conventional MRI variables reflecting lesion load (i.e., lesion count and lesion volume).

Finally, to further study the clinical usefulness of our approach, patients were stratified according to topology of their remaining connectivity and their lesion load. Global efficiency was chosen as topological variable of interest as previous studies proved its utility in clinical applications for MS (Fleischer et al., 2019; Rocca et al., 2016; Yaou et al., 2017). In particular, for every cohort, patients had low lesion load when their total lesion volume (TLV) was below the average TLV within their cohort. Similarly, patients had low, or high global efficiency with respect to the average value within each cohort. To investigate the complementarity and added value of global efficiency compared to TLV, we compared EDSS distributions among groups characterized by either similar TLV or GE levels using non-parametric Wilcoxon tests with post-hoc correction for multiple comparisons using Benjamini Hochberg’s method. The so-obtained p-values were further combined in a meta-analysis using weighted sum of z method (Zaykin, 2011) to investigate the consistency of our findings across the three cohorts.

## 4. Results

### 4.1. Comparing Disconnectome Models

The connection-level rank-rank Spearman correlation between atlas-based and individual tractography disconnectomes computed in patients from the diffusion cohort was 0.76±0.07 and 0.75±0.07 for automated and manual lesion segmentation respectively and is shown in Supplementary Figure 2.

#### 4.1.1. Large-scale Topological Features

Large-scale topological features extracted from atlas-based disconnectome graphs were plotted against equivalent features derived from individual tractography-based disconnectomes in Figure 4 for both manual segmentation (in yellow) and automated LeMan-PV segmentation (in blue) for the diffusion cohort. Intraclass correlation ICC(3,1) and Spearman correlation between large-scale atlas-based and individual tractography-based features in the diffusion cohort, extracted with both manual and automated LeMan-PV segmentations are reported together with significance level after Benjamini-Hochberg false discovery rate (FDR) correction. Overall, manual segmentation allowed a slightly stronger although similar correlation between atlas-based and individual tractography-based features. Spearman’s correlation was equal or above 0.73 for all features. ICC was higher for transitivity, average clustering, and average neighbour degree (>0.6). Comparatively small ICC (<0.5) were observed not only for average betweenness centrality, but also for global efficiency and average shortest path length.

**Figure 4.**
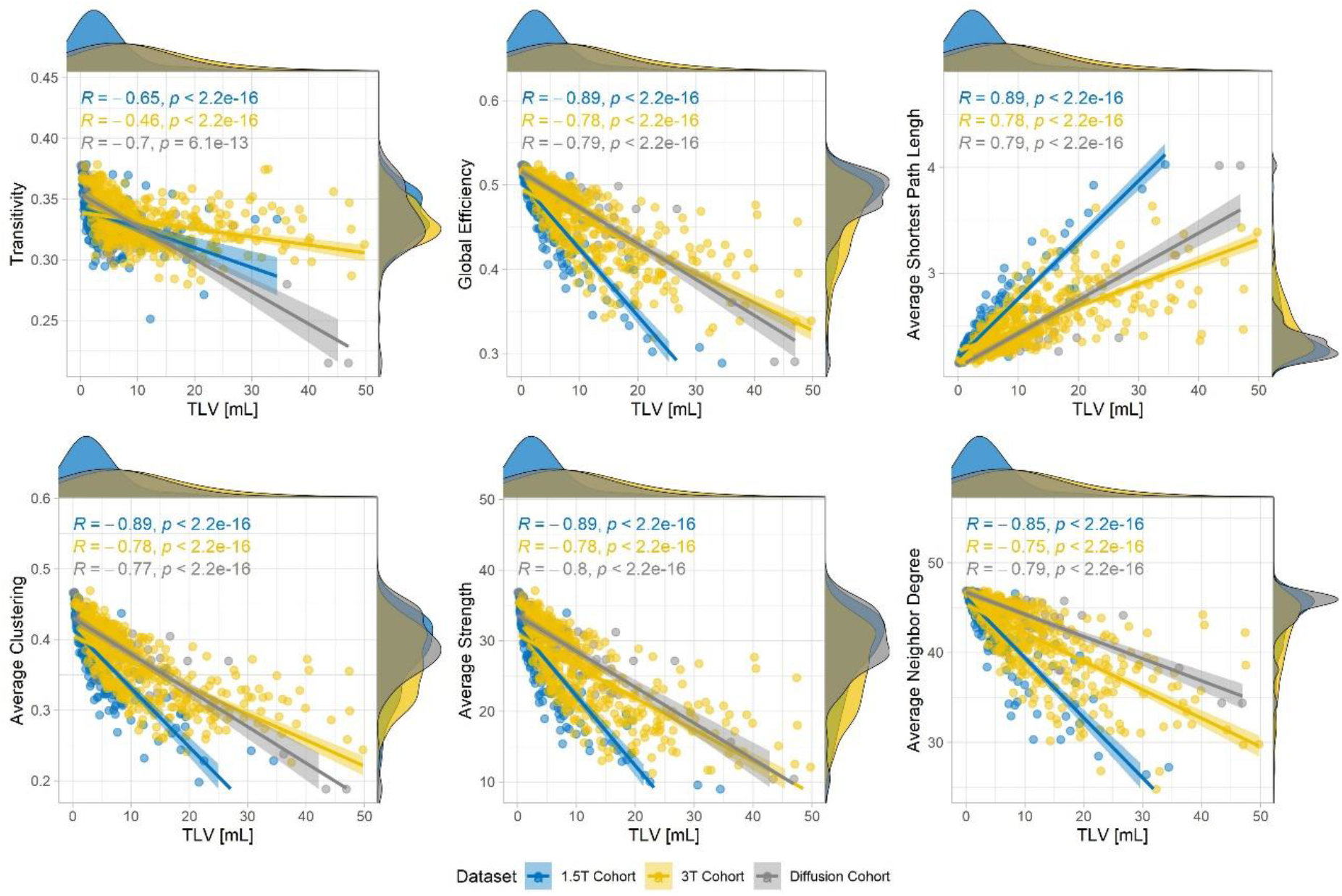
Variation of large scale topological features (y-axis) with total lesion volume (TLV, x-axis) for patients in all datasets (diffusion cohort in grey, 1.5T cohort in blue and 3T cohort in yellow). Spearman’s R and p-value are reported for all datasets.

#### 4.1.2. Small-scale Topological Features

Average correlations and standard deviation computed across all patients are reported in Table 3, together with combined p-values using Fisher’s method and respective χ^2^ statistics. Overall, the results show that atlas-based features are highly correlated with individual tractography-based features (ICC>0.75, ρ>0.70), except for the betweenness centrality (ICC<0.25, ρ<0.6), in good accordance with previous observations in Section 4.1.1. Further, automated lesion segmentation did not substantially impact the correlation between atlas-based and individual tractography-based features..

**Table 3.** Intraclass correlation and Spearman ρ between small-scale atlas-based and Individual tractography-based small-scale features and the associated p-value adjusted for FDR, for both manual and automated LeMan-PV lesion segmentation. The ICC is expressed as the mean over the population ± standard deviation. P-values were combined using Fisher’s method across the population. The combined p-value is reported together with the respective χ^2^ statistics

|  | Node strength |  | Neighbour degree |  | Node Clustering |  | Betweenness Centrality |  |
| --- | --- | --- | --- | --- | --- | --- | --- | --- |
| Segmentation | Manual | LeMan-PV | Manual | LeMan-PV | Manual | LeMan-PV | Manual | LeMan-PV |
| ICC | 0.85±0.16 | 0.76±0.2 | 0.90±0.08 | 0.86±0.12 | 0.87 ± 0.8 | 0.84 ± 0.08 | 0.20±0.1 | 0.19±0.08 |
| P | <10<sup>-16</sup> | <10<sup>-16</sup> | <10<sup>-16</sup> | <10<sup>-16</sup> | <10<sup>-16</sup> | <10<sup>-16</sup> | <10<sup>-16</sup> | <10<sup>-16</sup> |
| ($\chi^2_{icc}$) | (19550) | (14898) | (19905) | (17875) | (17905) | (15698) | (724) | (647) |
| $\rho$ | 0.86±0.14 | 0.78±0.18 | 0.90±0.09 | 0.85±0.15 | 0.76±0.14 | 0.70±0.16 | 0.54±0.12 | 0.52±0.10 |
| p | <10<sup>-16</sup> | <10<sup>-16</sup> | <10<sup>-16</sup> | <10<sup>-16</sup> | <10<sup>-16</sup> | <10<sup>-16</sup> | <10<sup>-16</sup> | <10<sup>-16</sup> |
| ($\chi^2_{\rho}$) | (19136) | (15024) | (20691) | (17192) | (11442) | (9350) | (4376) | (3973) |

Figure 5 shows how intraclass correlation and Spearman correlation vary with TLV estimated from either manual segmentation (in yellow) or LeMan-PV (in blue). Overall, the weakest correlations were observed for larger lesion loads and segmentation techniques did not substantially impact the correlation distributions.

**Figure 5.**
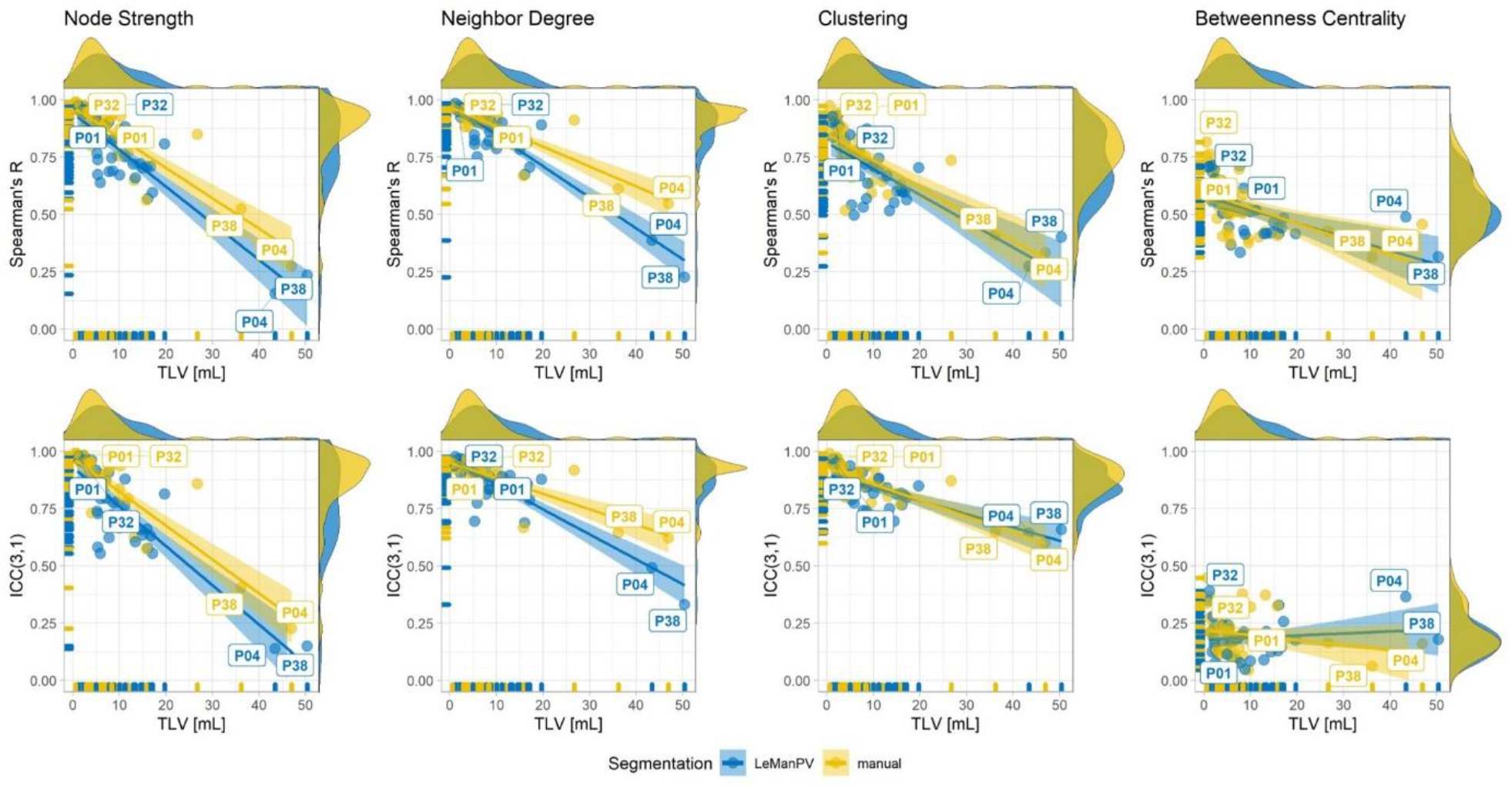
Variation of correlation of small-scale features extracted from individual versus atlas-based tractography with lesion load for the diffusion dataset. Each point is a patient. Spearman correlation and ICC(3,1) agreement between the two methods are computed across nodes for all patients and are shown on the y-axis. The total lesion volume (TLV) estimated from either manual (in yellow) or LeMan-PV lesion segmentations (in blue) on the x-axis. Labels in the graph indicate patient with generally high or low agreement, for which details are provided in Supplementary Figure 3.

**Figure 5.**
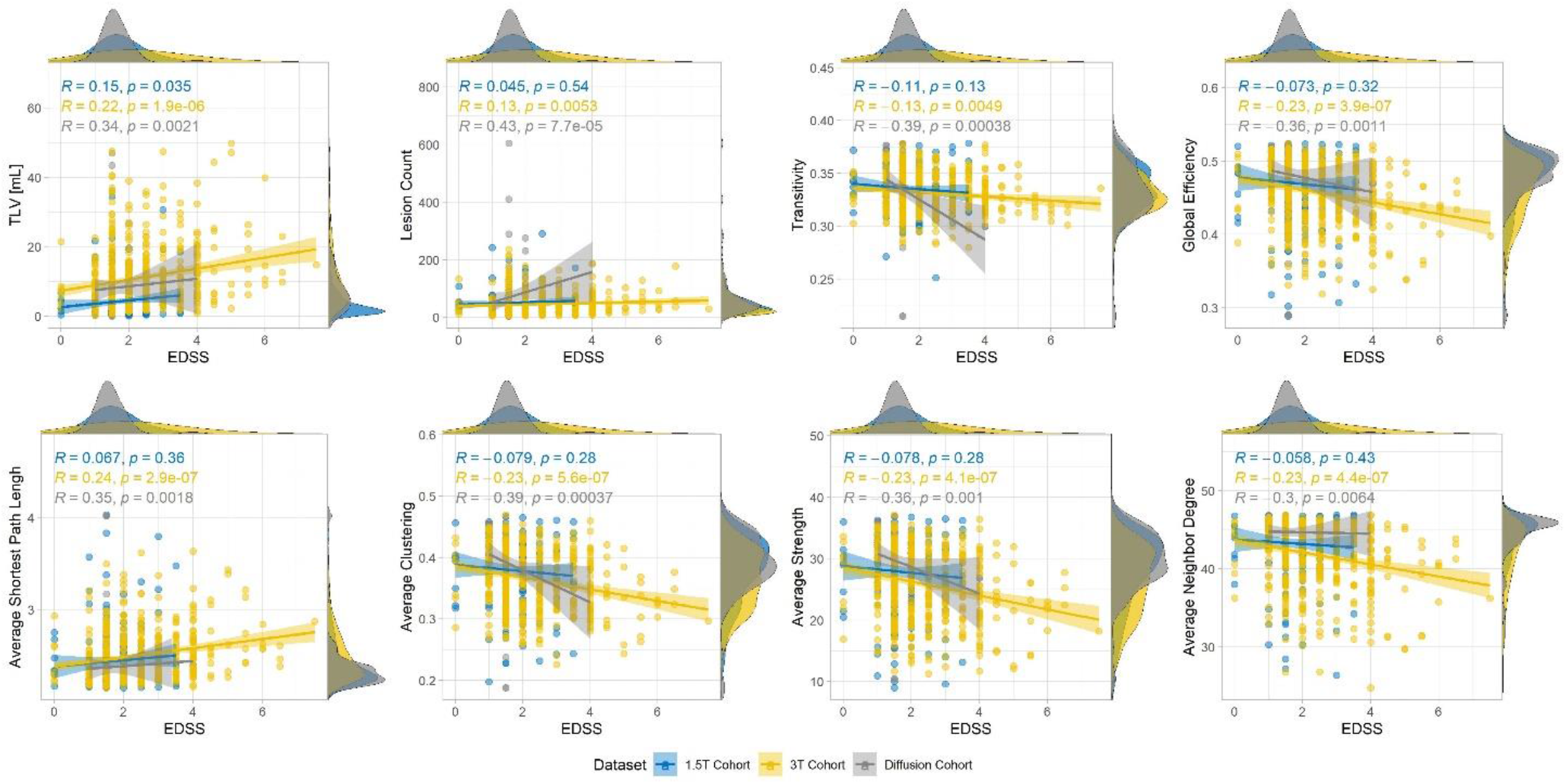
Variation of large scale topological features (y-axis) with Expanded Disability Status Scale (EDSS, x-axis) for patients in all datasets (diffusion cohort in grey, 1.5T cohort in blue and 3T cohort in yellow). Spearman’s R and p-value are reported for all datasets.

In Supplementary Figure 3 two patients with overall high correlations (P01 and P32) and low lesion load are shown together with two patients with overall lower correlations (P04 and P38) and higher lesion load.

Supplementary Table 2 reports the correlations between relative absolute differences in topology and age, brain volume and segmentation quality. Overall, the correlations were poor (R<=0.31), and the only significant finding was a negative correlation of global efficiency (GE) with manual lesion segmentation (R=-0-31, p=0.05). However, when corrected for multiple comparison, no significant correlation was found.

### 4.2. Clinical Applications

The variation of large-scale topological features with the TLV (estimated from manual segmentation for the diffusion and the 1.5T datasets and from LeMan-PV for the 3T dataset) is represented in Figure 6 for patients in the three datasets. As betweenness centrality was shown to be poorly estimated with our atlas-based approach, it was discarded from further analysis. Overall, patients in the 1.5T cohort had lower lesion load than other patients. All features showed strong and significant Spearman correlations with TLV in all datasets. The weakest correlations were observed for the transitivity, especially when computed for the 3T cohort (R=-0.46). Transitivity, global efficiency, average clustering, average strength, and average neighbour degree were found to decrease with increasing lesion load, whereas the average shortest path length increased.

**Figure 6.**
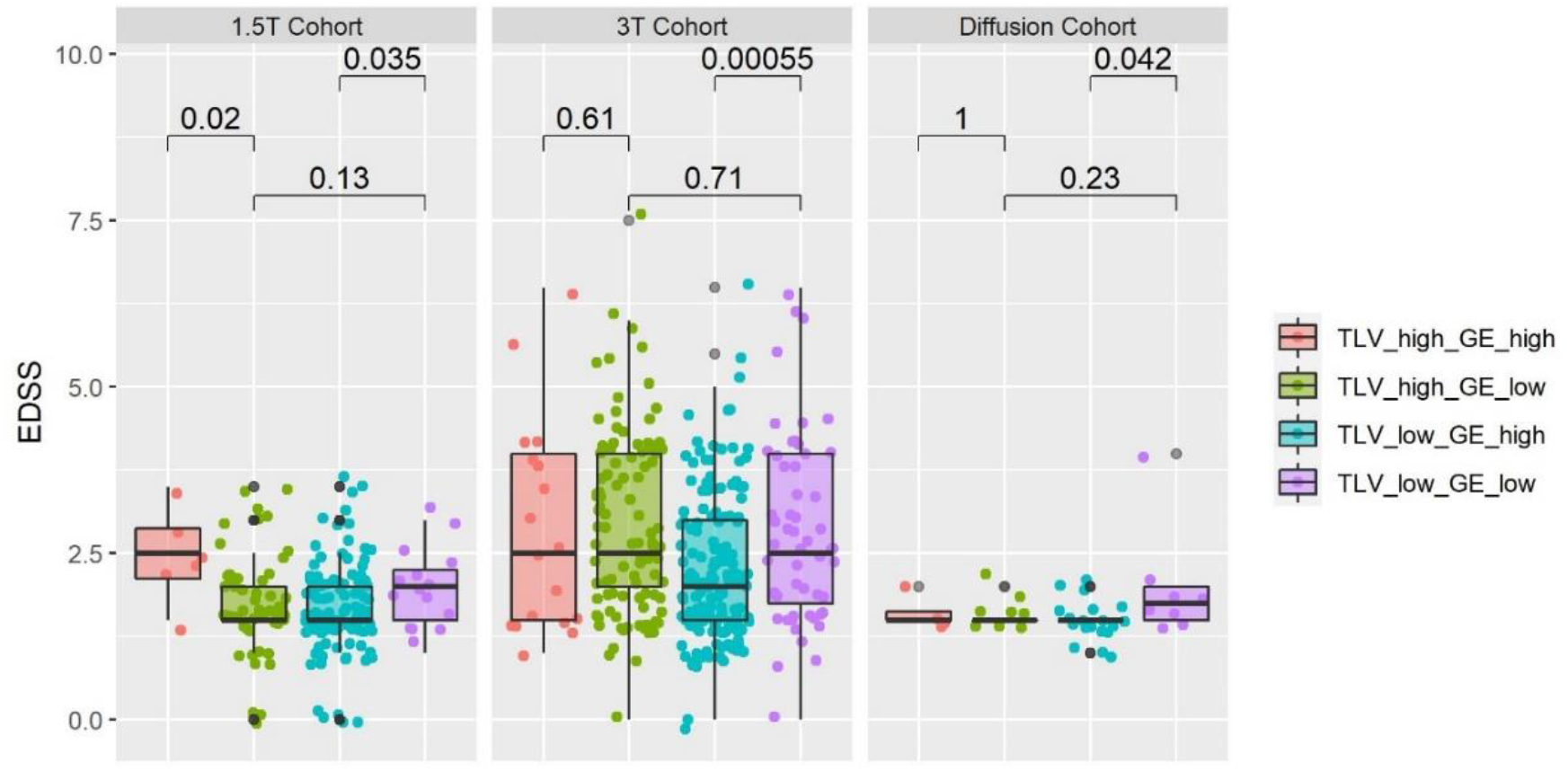
EDSS distribution in all cohorts when stratified according to total lesion volume (TLV) and the global efficiency (GE) of their remaining connectivity. The cut-offs used to classify patients were the average TLV and GE within each cohort.

The variation of large-scale features is also plotted against EDSS in Figure 7 for the three datasets. For comparison, the correlation between variables reflecting the lesion load (lesion count and TLV) and the EDSS are also reported. Patients in the 3T dataset had overall more severe and more heterogeneous EDSS than patients in the 1.5T and diffusion datasets. Lesion count and TLV were comparably correlated with EDSS for all datasets. The weakest correlations were consistently observed for the 1.5T dataset, whereas the findings were generally in good accordance between the 3T and the diffusion datasets. Overall, the correlation between our topological variables and EDSS was comparable but not substantially stronger than the correlations with TLV and lesion count.

When stratified according to TLV and global efficiency, patients consistently showed significantly different EDSS across datasets (see Figure 8). Adjusted p-values are reported in Table 4 for each cohort. The cut-off used to stratify patients according to their TLV and GE were 13.0 mL and 0.449 for the 3T cohort; 4.2 mL and 0.466 for the 1.5T cohort; and 7.7 mL and 0.475 for the diffusion cohort. In particular, among patients with low TLV, those who were also characterized by a low global efficiency had more severe EDSS than those with higher global efficiency in the 3T and 1.5T, whilst these findings were not significant when corrected for multiple comparisons in the diffusion cohort.

**Table 4.** Comparison of EDSS distributions among pairs of groups characterized by the same level of total lesion volume (TLV, first two rows) or global efficiency (GE, last two rows), computed for each cohort. The adjusted p-values using Benjamini Hochberg are reported together with the Wilcoxon statistics W. (*<0.05, **<0.01).

|  | 1.5T Cohort |  | 3T Cohort |  | Diffusion Cohort |  |
| --- | --- | --- | --- | --- | --- | --- |
|  | W | p-adjusted | W | p-adjusted | W | p-adjusted |
| **High TLV** | 244 | **0.041 \*** | 841 | 0.71 | 18.5 | 1 |
| **Low TLV** | 629 | **0.047 \*** | 3371 | **0.002 \*\*** | 42.5 | 0.17 |
| **High GE** | 602 | **0.026 \*** | 1964 | 0.175 | 46 | 0.60 |
| **Low GE** | 292 | 0.127 | 2878 | 0.71 | 25 | 0.46 |

These findings were significant when combined in a meta-analysis (sumz=3.18, p=7*10^™4^).

## 5. Discussion

In this work, we validated a novel approach to model structural disconnectomes without diffusion imaging in a large retrospective multi-centric study of multiple sclerosis. Similarly to what has been described in (Griffis et al., 2021), the HCP842 tractography atlas (Yeh et al., 2018) was used to extract brain connections transected by lesions, and the remaining connectivity was modelled as a brain graph. Disconnectome topological features were proposed as possible new imaging biomarkers.

Compared to previous studies that focused on the clinical impact of lesion location either in voxel-based regions (so called “lesion-symptom mapping” (Bates et al., 2003)) or within predefined brain networks (“lesion network mapping” (Fox, 2018)), our method allows to quantify the impact of lesions on the whole structural connectome.

When comparing atlas-based to individual tractography-based disconnectomes in MS patients, one should take into consideration that although the same reconstruction and tracking algorithms were used as for the tractography atlas, individual tractograms lacked the filtering of false positives which was performed manually during the construction of the atlas. Furthermore, our disconnectome method assumes that streamlines affected by lesions can be isolated from the structural connectome. However, individual tractography can already be influenced by the presence of lesions itself (Lin et al., 2005; Reich et al., 2007; Tievsky et al., 1999) and might not therefore reflect the actual structural connectivity of these patients.

The connection-level rank correlation between atlas-based and individual tractography-based models suggests that the strength of connections is overall consistently reflecting a substantial similarity between disconnectome graphs. Small-scale and large-scale topological features extracted from atlas-based disconnectomes and classical individual models were strongly correlated even though an offset was often observed, resulting in lower ICC. Despite the lack of absolute agreement, the consistency of these findings suggest that our atlas-based model is a well-suited approximation of individual connectivity loss for clinical applications. Nevertheless, the observed offset between individual tractography and atlas-based models should be taken in consideration when quantitively comparing these results with literature.

Further, for small-scale features, patients associated with low agreement between individual and atlas-based disconnectome models were found to be characterized by large TLV. This could be explained either by the interference of lesions with individual tractography, or by an inadequate approximation using the tractography atlas.

In terms of comparability between atlas-based and individual tractography-based methods, betweenness centrality was the worst performing feature. This observation suggests that our atlas-based method reflects well global or node properties that are independent from specific paths, whereas it does not allow a precise approximation of node neighbourhood. This could be explained by normal individual variability in axonal connections and by the erroneous or biased estimation of disconnectomes from individual tractography.

Using an automated lesion segmentation did not negatively impact the results, suggesting that a fully automated approach could be used to estimate disconnectome graphs using LeMan-PV segmentation estimated solely from T1-weighted and FLAIR contrasts. Further, the validation analysis was found to be independent from patients’ age, brain atrophy and segmentation quality.

Large-scale atlas-based topological features were shown to be significantly correlated with total lesion volume in all datasets. Importantly, the association between TLV and topological features was assumed to be linear, but other distributions could be better suited to reflect this correlation, especially for transitivity and average clustering which seem to follow a logarithmic function. These observations were coherent for all datasets despite substantial differences in terms of demographics, lesion load and clinical scores. However, different brain parcellation techniques might influence the graph properties due to different definitions of a “node” and should be explored in future work. In general, a greater variability is expected to be induced by smaller brain regions, leading to a bias-variance trade-off.

In multiple sclerosis patients, values decreased for increasing lesion load for transitivity, average clustering, average strength, and global efficiency, whereas the average shortest path length increased, resulting in decreased small-worldness. This is consistent with previous studies (Aerts et al., 2016; Faivre et al., 2016; Rocca et al., 2016, 2015) that showed significantly lower node strength, efficiency, clustering and increased shortest path length in MS patients when compared to controls (Shu et al., 2016), and impaired small-world efficiency associated to white matter lesion load. Interestingly, the distribution of topological measures were closer between the 3T and diffusion cohorts despite different lesion segmentation strategies. Such observation could be explained by the different lesion load distribution of the 1.5T cohort and the substantially different acquisition protocols.

Univariate correlations between large-scale topological features and EDSS were generally poor but close to lesion load variables. Previous studies showed a significant decrease in global efficiency although weakly correlated with EDSS (Muthuraman et al., 2016; Shu et al., 2011). Weaker correlations were observed for the 1.5T dataset, probably due to the narrower distribution of EDSS values.

When stratified according to their TLV and global efficiency, patients with low lesion load showed significantly different EDSS depending on their global efficiency value: the lower the global efficiency, the worst the clinical outcome. These findings were validated across all cohorts and suggest that disconnectome topological features could be used in addition to TLV to stratify patients into subgroups reflecting different clinical states. In patients with high TLV, significantly higher EDSS values were observed for higher global efficiency in the 1.5T cohort. A high remyelination capacity might explain these results since the 1.5T cohort was composed by patients at very early stages of the disease. Another hypothesis to explain these findings is the interplay of other pathophysiological effects we are not considering, such as the influence of spinal cord and optic nerve lesions on EDSS. When interpreting these stratification results, one should note that the strong correlation between GE and TLV results into unbalanced groups, therefore possibly influencing the statistical analysis. Further, demographics and disease-related features (e.g., disease duration) were not considered for the stratification and might also explain the observed difference in EDSS distribution.

Due to the limited age range covered by the tractography atlas, we implemented an applicability criterion that restricts the target clinical population to patients without profound atrophy. Therefore, the findings of this work must be interpreted accordingly and cannot be extrapolated to all multiple sclerosis patients and in particular to aging populations or cases with extensive atrophy. However, our method is suitable for conditions such as stroke, TBI and MS, which occur with high prevalence in mid-aged adults. In addition to global brain atrophy, regional atrophy levels might also impact the anatomy of neuronal connections and should be deeper investigated in future work. Also, the contribution of brain plasticity which might yield to a partial or complete recovery of neuronal connections (Aerts et al., 2016; Fleischer et al., 2019; Rocca et al., 2015) is not considered in our atlas-based approach and could impact the correlation with clinical scores. Finally, even though the use of a tractography atlas mitigates intra-subject variability related to the diffusion acquisition and tractography algorithms, other commonly known pitfalls of brain tractography remain, such as the dependence on the modelling procedure of the axonal direction in the voxels. Therefore, the results might vary depending on the tractography atlas used based on the underlying diffusion data and processing.

Different types of lesions might differently impact the connectivity loss based on their active or inactive state, as well as ongoing remyelination and demyelination processes. Future work should aim at investigating this relation using contrast enhanced scans to identify active lesions, whilst T1 hypointensities could be used to find “black holes” (Truyen et al., 1996).

Another limitation of our approach is that it only considers connectivity changes in response to focal WM lesions. However, WM lesions are only a part of the complex pathophysiology of MS, and other mechanisms might also influence connectivity impairments, such as microstructural damage in normal appearing white matter (Solana et al., 2018). Furthermore, the extent of connectivity loss caused by a T2-weighted hyperintense lesion was not explored in our work, as streamlines were tagged either as affected (when intersecting the lesion mask) or not affected. A microstructural analysis of tissue properties within lesions (e.g., DTI-derived metrics) would allow a more precise estimation of connectivity damage. Whilst diffusion properties cannot be estimated with our tractography atlas-based approach, the combination of our disconnectome model with other quantitative MRI modalities is an interesting line of research for future work.

Last but not least, lesions located in the spine, cortex and optical nerve were not considered in this study but might play a role in explaining the clinico-radiological paradox in multiple sclerosis.

Previous studies have shown how brain connectivity and network efficiency in particular can be affected by the sex and brain size of the subject (Yan et al., 2011). Such effects are not considered in our approach as the tractography atlas is independent from the sex and brain size, which might therefore result into interpretation pitfalls.

## 6. Conclusion

In conclusion, our atlas-based graph model of disconnectivity allows to extract topological features shown to reflect actual physiological properties and in good accordance with individual tractography disconnectome models in multiple sclerosis. Such metrics were shown to contribute to narrowing the clinico-radiological gap in multiple sclerosis by providing a new quantitative characterization of brain disconnectivity, in a large retrospective multi-centric study involving substantially different clinical cohorts and MR images in terms of hardware and acquisition sequences.

This method could be applied to any other neurological diseases characterized by the appearance of white matter lesions to study their impact on structural connectivity without requiring diffusion imaging. This is especially interesting for clinical research, as our method enables the retrospective analysis of structural disconnectivity in pre-existing datasets with routine MR contrasts that does not contain high quality diffusion imaging.

## Data Availability

Due to patient privacy, the clinical data used in this study cannot be made openly available.

## Acknowledgments

We thank Frank Yeh, author of the tractography atlas and developer of DSI Studio, for the fruitful discussion and his advice on the reconstruction and tracking algorithms used. We also thank Dimitri Van De Ville for gracefully hosting this project and providing useful feedback and discussion.

## Author Contributions

**Veronica Ravano:** Conceptualization, Methodology, Software, Formal analysis, Investigation, Writing – Original draft preparation, Visualization; **Michaela Andelova:** Data curation, Investigation, Resources; **Mário João Fartaria**: Software, Data curation; **Mazen Fouad A-Wali Mahdi:** Software; **Reto Meuli**: Funding acquisition, Resources; **Bénédicte Maréchal:** Software, Data curation ; **Tomas Uher**: Data curation, Resources; **Jan Krasensky:** Data curation, Resources; **Manuela Vaneckova:** Data curation, Funding acquisition, Resources; **Dana Horakova:** Data curation, Funding acquisition, Resources; **Tobias Kober:** Conceptualization, Funding acquisition, Supervision; **Jonas Richiardi:** Conceptualization, Methodology, Investigation, Writing – Original draft preparation, Supervision, Project administration; **all:** Writing – Review and Editing

## References

Aerts, H., Fias, W., Caeyenberghs, K., Marinazzo, D., 2016. Brain networks under attack: Robustness properties and the impact of lesions. Brain 139, 3063–3083. https://doi.org/10.1093/brain/aww194

Barkhof, F., 2002. The clinico-radiological paradox in multiple sclerosis revisited. Current Opinion in Neurology 15, 239–245. https://doi.org/10.1097/00019052-200206000-00003

Bassett, D.S., Sporns, O., 2017. Network neuroscience. NATURE NEUROSCIENCE 20, 353–364. https://doi.org/10.1038/nn.4502

Bates, E., Wilson, S.M., Saygin, A.P., Dick, F., Sereno, M.I., Knight, R.T., Dronker, N.F., 2003. Voxel-based lesion-symptom mapping. Nature Neuroscience 6, 448–450. https://doi.org/10.1038/nn1050

Buckner, R.L., Krienen, F.M., Castellanos, A., Diaz, J.C., Yeo, B.T.T., 2011. The organization of the human cerebellum estimated by intrinsic functional connectivity. J Neurophysiol 106, 2322–2345. https://doi.org/10.1152/jn.00339.2011.

Bullmore, E.T., Bassett, D.S., 2011. Brain Graphs: Graphical Models of the Human Brain Connectome. Annual Review of Clinical Psychology 7, 113–140. https://doi.org/10.1146/annurev-clinpsy-040510-143934

Bullmore, E.T., Sporns, O., 2009. Complex brain networks : graph theoretical analysis of structural and functional systems. Nature Reviews Neuroscience 10, 186–198. https://doi.org/10.1038/nrn2575

Butzkueven, H., Chapman, J., Cristiano, E., Grand’Maison, F., Hoffmann, M., Izquierdo, G., Jolley, D., Kappos, L., Leist, T., Pöhlau, D., Rivera, V., Trojano, M., Verheul, F., Malkowski, J.P., 2006. MSBase: An international, online registry and platform for collaborative outcomes research in multiple sclerosis. Multiple Sclerosis 12, 769–774. https://doi.org/10.1177/1352458506070775

Calabrese, E., Badea, A., Coe, C.L., Lubach, G.R., Styner, M.A., Johnson, G.A., 2014. Investigating the tradeoffs between spatial resolution and diffusion sampling for brain mapping with diffusion tractography: Time well spent? Human Brain Mapping 35, 5667–5685. https://doi.org/10.1002/hbm.22578

Carrera, E., Tononi, G., 2014. Diaschisis: Past, present, future. Brain 137, 2408–2422. https://doi.org/10.1093/brain/awu101

Catani, M., Dell’Acqua, F., Bizzi, A., Forkel, S.J., Williams, S.C., Simmons, A., Murphy, D.G., Thiebaut de Schotten, M., 2012. Beyond cortical localization in clinico-anatomical correlation. Cortex 48, 1262–1287. https://doi.org/10.1016/j.cortex.2012.07.001

Catani, M., Ffytche, D.H., 2005. The rises and falls of disconnection syndromes. Brain 128, 2224–2239. https://doi.org/10.1093/brain/awh622

Charil, A., Zijdenbos, A.P., Taylor, J., Boelman, C., Worsley, K.J., Evans, A.C., Dagher, A., 2003. Statistical mapping analysis of lesion location and neurological disability in multiple sclerosis : application to 452 patient data sets. NeuroImage 19, 532–544. https://doi.org/10.1016/S1053-8119(03)00117-4

Chow, L.S., Paramesran, R., 2016. Review of medical image quality assessment. Biomedical Signal Processing and Control 27, 145–154. https://doi.org/10.1016/j.bspc.2016.02.006

Ciccarelli, O., Catani, M., Johansen-Berg, H., Clark, C., Thompson, A., 2008. Diffusion-based tractography in neurological disorders: concepts, applications, and future developments. The Lancet Neurology 7, 715–727. https://doi.org/10.1016/S1474-4422(08)70163-7

Connors, J., Krzywinski, M., Schein, J., Gascoyne, R., Horsman, D., Jones, S.J., Marra, M.A., 2009. Circos : An information aesthetic for comparative genomics. Genome Research 19, 1639–1645. https://doi.org/10.1101/gr.092759.109.19

De Vico Fallani, F., Richiardi, J., Chavez, M., Achard, S., 2014. Graph analysis of functional brain networks: Practical issues in translational neuroscience. Philosophical Transactions of the Royal Society B 369. https://doi.org/10.1098/rstb.2013.0521

Donahue, C.J., Sotiropoulos, S.N., Jbabdi, S., Hernandez-Fernandez, M., Behrens, T.E., Dyrby, T.B., Coalson, T., Kennedy, H., Knoblauch, K., Van Essen, D.C., Glasser, M.F., 2016. Using diffusion tractography to predict cortical connection strength and distance: A quantitative comparison with tracers in the monkey. Journal of Neuroscience 36, 6758–6770. https://doi.org/10.1523/JNEUROSCI.0493-16.2016

Dziedzic, T., Metz, I., Dallenga, T., König, F.B., Müller, S., Stadelmann, C., Brück, W., 2010. Wallerian degeneration: A major component of early axonal pathology in multiple sclerosis. Brain Pathology 20, 976–985. https://doi.org/10.1111/j.1750-3639.2010.00401.x

Faivre, A., Robinet, E., Guye, M., Rousseau, C., Maarouf, A., Le Troter, A., Zaaraoui, W., Rico, A., Crespy, L., Soulier, E., Confort-Gouny, S., Pelletier, J., Achard, S., Ranjeva, J.P., Audoin, B., 2016. Depletion of brain functional connectivity enhancement leads to disability progression in multiple sclerosis: A longitudinal resting-state fMRI study. Multiple Sclerosis 22, 1695–1708. https://doi.org/10.1177/1352458516628657

Fan, L., Li, H., Zhuo, J., Zhang, Y., Wang, J., Chen, L., Yang, Z., Chu, C., Xie, S., Laird, A.R., Fox, P.T., Eickhoff, S.B., Yu, C., Jiang, T., 2016. The Human Brainnetome Atlas: A New Brain Atlas Based on Connectional Architecture. Cerebral Cortex 26, 3508–3526. https://doi.org/10.1093/cercor/bhw157

Fartaria, M.J., Bonnier, G., Roche, A., Kober, T., Meuli, R., Rotzinger, D., Frackowiak, R., Schluep, M., Du Pasquier, R., Thiran, J.P., Krueger, G., Bach Cuadra, M., Granziera, C., 2016. Automated detection of white matter and cortical lesions in early stages of multiple sclerosis. Journal of Magnetic Resonance Imaging 43, 1445–1454. https://doi.org/10.1002/jmri.25095

Fartaria, M.J., Roche, A., Meuli, R., Granziera, C., Kober, T., Bach Cuadra, M., 2017. Segmentation of Cortical and Subcortical Multiple Sclerosis Lesions Based on Constrained Partial. MICCAI LCNS 10435, 516–524. https://doi.org/10.1007/978-3-319-66179-7

Fleischer, V., Radetz, A., Ciolac, D., Muthuraman, M., Gonzalez-escamilla, G., 2019. Graph Theoretical Framework of Brain Networks in Multiple Sclerosis : A Review of Concepts. Neuroscience 403, 35–53. https://doi.org/10.1016/j.neuroscience.2017.10.033

Foulon, C., Cerliani, L., Kinkingnéhun, S., Levy, R., Rosso, C., Urbanski, M., Volle, E., de Schotten, M.T., 2018. Advanced lesion symptom mapping analyses and implementation as BCBtoolkit. GigaScience 7, 1–17. https://doi.org/10.1093/gigascience/giy004

Fox, M., 2018. Mapping Symptoms to Brain Networks with the Human Connectome. The New England Journal of Medicine 379, 2237–2245. https://doi.org/10.1056/NEJMra1706158

Gorgoraptis, N., Wheeler-Kingshott, C.A.M., Jenkins, T.M., Altmann, D.R., Miller, D.H., Thompson, A.J., Ciccarelli, O., 2010. Combining tractography and cortical measures to test system-specific hypotheses in multiple sclerosis. Multiple Sclerosis 16, 555–565. https://doi.org/10.1177/1352458510362440

Griffis, J.C., Metcalf, N. v., Corbetta, M., Shulman, G.L., 2021. Lesion Quantification Toolkit: A MATLAB software tool for estimating grey matter damage and white matter disconnections in patients with focal brain lesions. NeuroImage: Clinical 30, 102639. https://doi.org/10.1016/j.nicl.2021.102639

Hagberg, A.A., Schult, D.A., Swart, P.J., 2008. Exploring network structure, dynamics, and function using NetworkX. 7th Python in Science Conference (SciPy 2008) 11–15.

Hagmann, P., Cammoun, L., Gigandet, X., Meuli, R., Honey, C.J., Van Wedeen, J., Sporns, O., 2008. Mapping the structural core of human cerebral cortex. PLoS Biology 6, 1479–1493. https://doi.org/10.1371/journal.pbio.0060159

Hamilton, W.L., Ying, R., Leskovec, J., 2017. Representation Learning on Graphs: Methods and Applications. Bulletin of the IEEE Computer Society Technical Committee on Data Engineering 1–24.

Hayes, C.E., Ntambi, J.M., 2020. Multiple Sclerosis: Lipids, Lymphocytes, and Vitamin D. Immunometabolism 1–54. https://doi.org/10.20900/immunometab20200019

Horakova, D., Zivadinov, R., Weinstock-Guttman, B., Havrdova, E., Qu, J., Tamaño-Blanco, M., Badgett, D., Tyblova, M., Bergsland, N., Hussein, S., Willis, L., Krasensky, J., Vaneckova, M., Seidl, Z., Lelkova, P., Dwyer, M.G., Zhang, M., Yu, H., Duan, X., Kalincik, T., Ramanathan, M., 2013. Environmental Factors Associated with Disease Progression after the First Demyelinating Event: Results from the Multi-Center SET Study. PLoS ONE 8, 1–8. https://doi.org/10.1371/journal.pone.0053996

Horn, T., Sherwood, J., Remien, R.H., Nash, D., Auerbach, J.D., 2016. Towards an integrated primary and secondary HIV prevention continuum for the United States: A cyclical process model. J. Int. AIDS Soc. 19, 1–6. https://doi.org/10.7448/IAS.19.1.21263

Jones, D.K., Knösche, T.R., Turner, R., 2013. White matter integrity, fiber count, and other fallacies: The do’s and don’ts of diffusion MRI. NeuroImage 73, 239–254. https://doi.org/10.1016/j.neuroimage.2012.06.081

Klein, S., Staring, M., Murphy, K., Viergever, M. a., Pluim, J., 2010. elastix: A Toolbox for Intensity-Based Medical Image Registration. IEEE Transactions on Medical Imaging 29, 196–205. https://doi.org/10.1109/TMI.2009.2035616

Kuceyeski, A.F., Vargas, W., Dayan, M., Monohan, E., Blackwell, C., Raj, A., Fujimoto, K., Gauthier, S.A., 2015. Modeling the relationship among gray matter atrophy, abnormalities in connecting white matter, and cognitive performance in early multiple sclerosis. American Journal of Neuroradiology 36, 702–709. https://doi.org/10.3174/ajnr.A4165

Lin, X., Tench, C.R., Morgan, P.S., Niepel, G., Constantinescu, C.S., 2005. “Importance sampling” in MS: Use of diffusion tensor tractography to quantify pathology related to specific impairment. Journal of the Neurological Sciences 237, 13–19. https://doi.org/10.1016/j.jns.2005.04.019

Lipp, I., Parker, G.D., Tallantyre, E.C., Goodall, A., Grama, S., Patitucci, E., Heveron, P., Tomassini, V., Jones, D.K., 2020. Tractography in the presence of multiple sclerosis lesions. NeuroImage 209, 116471. https://doi.org/10.1016/j.neuroimage.2019.116471

Llufriu, S., Martinez-Heras, E., Solana, E., Sola-Valls, N., Sepulveda, M., Blanco, Y., Martinez-Lapiscina, E.H., Andorra, M., Villoslada, P., Prats-Galino, A., Saiz, A., 2017. Structural networks involved in attention and executive functions in multiple sclerosis. NeuroImage: Clinical 13, 288–296. https://doi.org/10.1016/j.nicl.2016.11.026

Lucchinetti, C., Brück, W., Parisi, J., Scheithauer, B., Rodriguez, M., Lassmann, H., 2000. Heterogeneity of multiple sclerosis lesions: Implications for the pathogenesis of demyelination. Annals of Neurology 47, 707–717. https://doi.org/10.1002/1531-8249(200006)47:6<707::AID-ANA3>3.0.CO;2-Q

Meskaldji, D.E., Fischi-Gomez, E., Griffa, A., Hagmann, P., Morgenthaler, S., Thiran, J.P., 2013. Comparing connectomes across subjects and populations at different scales. Neuroimage 80, 416–425. https://doi.org/10.1016/j.neuroimage.2013.04.084

Muthuraman, M., Fleischer, V., Kolber, P., Luessi, F., Zipp, F., Groppa, S., 2016. Structural brain network characteristics can differentiate CIS from early RRMS. Frontiers in Neuroscience 10, 1–12. https://doi.org/10.3389/fnins.2016.00014

Pawlitzki, M., Neumann, J., Kaufmann, J., Heidel, J., Stadler, E., Sweeney-Reed, C., Sailer, M., Schreiber, S., 2017. Loss of corticospinal tract integrity in early MS disease stages. Neurology: Neuroimmunology and NeuroInflammation 4. https://doi.org/10.1212/NXI.0000000000000399

Ravano, V., Andelova, M., Fouad, M., Mahdi, A.-W., Meuli, R., Uher, T., Krasensky, J., Vaneckova, M., Horakova, D., Kober, T., Richiardi, J., 2020. Automated atlas-based mapping of white matter tract damage to multiple sclerosis symptoms, in: Proceedings of the International Society of Magnetic Resonance in Medicine. p. 1391.

Ravano, V., Andelova, Michaela Mahdi M.F.A.-W., Meuli, R., Uher, T., Krasensky, J., Vaneckova, M., Horakova, D., Kober, T., Richiardi, J., 2019. Atlas-based tract damage mapping improves 4-year forecast of EDSS in multiple sclerosis, in: Multiple Sclerosis Journal, Vol. 25. pp. 182–183.

Reich, D.S., Smith, S.A., Zackowski, K.M., Gordon-Lipkin, E.M., Jones, C.K., Farrell, J.A.D., Mori, S., van Zijl, P.C.M., Calabresi, P.A., 2007. Multiparametric magnetic resonance imaging analysis of the corticospinal tract in multiple sclerosis. NeuroImage 38, 271–279. https://doi.org/10.1016/j.neuroimage.2007.07.049

Richiardi, J., Achard, S., Bunke, H., Ville, D. Van De, 2013. Machine Learning with Brain Graphs. IEEE SIGNAL PROCESSING MAGAZINE 30, 58–70. https://doi.org/10.1109/MSP.2012.2233865

Rocca, M.A., Amato, M.P., De Stefano, N., Enzinger, C., Geurts, J.J., Penner, I.K., Rovira, A., Sumowski, J.F., Valsasina, P., Filippi, M., 2015. Clinical and imaging assessment of cognitive dysfunction in multiple sclerosis. The Lancet Neurology 14, 302–317. https://doi.org/10.1016/S1474-4422(14)70250-9

Rocca, M.A., Valsasina, P., Meani, A., Falini, A., Comi, G., Filippi, M., 2016. Impaired functional integration in multiple sclerosis: a graph theory study. Brain Structure and Function 221, 115–131. https://doi.org/10.1007/s00429-014-0896-4

Schmitter, D., Roche, A., Maréchal, B., Ribes, D., Abdulkadir, A., Bach-Cuadra, M., Daducci, A., Granziera, C., Klöppel, S., Maeder, P., Meuli, R., Krueger, G., 2015. An evaluation of volume-based morphometry for prediction of mild cognitive impairment and Alzheimer’s disease. NeuroImage: Clinical 7, 7–17. https://doi.org/10.1016/j.nicl.2014.11.001

Shu, N., Duan, Y., Xia, M., Schoonheim, M.M., Huang, J., Ren, Z., Sun, Z., Ye, J., Dong, H., Shi, F.D., Barkhof, F., Li, K., Liu, Y., 2016. Disrupted topological organization of structural and functional brain connectomes in clinically isolated syndrome and multiple sclerosis. Scientific Reports 6, 1–11. https://doi.org/10.1038/srep29383

Shu, N., Liu, Y., Li, K., Duan, Y., Wang, J., Yu, C., Dong, H., Ye, J., He, Y., 2011. Diffusion tensor tractography reveals disrupted topological efficiency in white matter structural networks in multiple sclerosis. Cerebral Cortex 21, 2565–2577. https://doi.org/10.1093/cercor/bhr039

Simioni, S., Amarù, F., Bonnier, G., Kober, T., Rotzinger, D., Du Pasquier, R., Schluep, M., Meuli, R., Sbarbati, A., Thiran, J.P., Krueger, G., Granziera, C., 2014. MP2RAGE provides new clinically-compatible correlates of mild cognitive deficits in relapsing-remitting multiple sclerosis. Journal of Neurology 261, 1606–1613. https://doi.org/10.1007/s00415-014-7398-4

Solana, E., Martinez-Heras, E., Martinez-Lapiscina, E.H., Sepulveda, M., Sola-Valls, N., Bargalló, N., Berenguer, J., Blanco, Y., Andorra, M., Pulido-Valdeolivas, I., Zubizarreta, I., Saiz, A., Llufriu, S., 2018. Magnetic resonance markers of tissue damage related to connectivity disruption in multiple sclerosis. NeuroImage: Clinical 20, 161–168. https://doi.org/10.1016/j.nicl.2018.07.012

Sporns, O., Tononi, G., Kötter, R., 2005. The human connectome: A structural description of the human brain. PLoS Computational Biology 1, 0245–0251. https://doi.org/10.1371/journal.pcbi.0010042

Telesford, Q.K., Joyce, K.E., Hayasaka, S., Burdette, J.H., Laurienti, P.J., 2011. The Ubiquity of Small-World Networks. Brain Connectivity 1, 367–375. https://doi.org/10.1089/brain.2011.0038

Tievsky, A.L., Ptak, T., Farkas, J., 1999. Investigation of apparent diffusion coefficient and diffusion tensor anisotropy in acute and chronic multiple sclerosis lesions. American Journal of Neuroradiology 20, 1491–1499.

Truyen, L., Van Waesberghe, J.H.T.M., Van Walderveen, M.A.A., Van Oosten, B.W., Polman, C.H., Hommes, O.R., Adèr, H.J.A., Barkhof, F., 1996. Accumulation of hypointense lesions (‘black holes’) on T1 spin-echo MRI correlates with disease progression in multiple sclerosis. Neurology 47, 1469–1476.

Van Essen, D.C., Ugurbil, K., Auerbach, E., Barch, D., Behrens, T.E.J., Bucholz, R., Chang, A., Chen, L., Corbetta, M., Curtiss, S.W., Della Penna, S., Feinberg, D., Glasser, M.F., Harel, N., Heathj, A.C., Larson-Prior, L., Marcus, D., Michalareas, G., Moeller, S., Oostenveld, R., Petersen, S.E., Prior, F., Schlaggar, B.L., Smith, S.M., Snyder, A.Z., Xu, J., Yacoub, E., Consortium, W.-M.H., 2012. The Human Connectome Project: A data acquisition perspective. NeuroImage 62, 2222–2231. https://doi.org/10.1115/JRC2014-3865

Vellinga, M.M., Geurts, J.J.G., Rostrup, E., Uitdehaag, B.M.J., Polman, C.H., Barkhof, F., Vrenken, H., 2009. Clinical correlations of brain lesion distribution in multiple sclerosis. Journal of Magnetic Resonance Imaging 29, 768–773. https://doi.org/10.1002/jmri.21679

Watts, D.J., Strogatz, S.H., 1998. Small social network dynamic set. Science 393, 440–442.

Yan, C., Gong, G., Wang, J., Wang, D., Liu, D., Zhu, C., Chen, Z.J., Evans, A., Zang, Y., He, Y., 2011. Sex-and brain size-related small-world structural cortical networks in young adults: A DTI tractography study. Cerebral Cortex 21, 449–458. https://doi.org/10.1093/cercor/bhq111

Yaou, L., Hao, W., Yunyun, D., Jing, H., Zhuoqiong, R., Jing, Y., Huiqing, D., Fudong, S., Kuncheng, L., Jinhui, W., 2017. Functional Brain Network Alterations in Clinically Isolated Syndrome and Multiple Sclerosis: A Graph-based Connectome Study. Radiology 282, 534–541. https://doi.org/10.1148/radiol.2016152843

Yeh, F.C., Panesar, S., Fernandes, D., Meola, A., Yoshino, M., Fernandez-Miranda, J.C., Vettel, J.M., Verstynen, T., 2018. Population-averaged atlas of the macroscale human structural connectome and its network topology. NeuroImage 178, 57–68. https://doi.org/10.1016/j.neuroimage.2018.05.027

Yeh, F.C., Tseng, W.Y.I., 2011. NTU-90: A high angular resolution brain atlas constructed by q-space diffeomorphic reconstruction. NeuroImage 58, 91–99. https://doi.org/10.1016/j.neuroimage.2011.06.021

Zaykin, D. V., 2011. Optimally weighted Z-test is a powerful method for combining probabilities in meta-analysis. J. Evol. Biol. 24, 1836–1841. https://doi.org/10.1111/j.1420-9101.2011.02297.x

