## Supplementary Material for "Validating atlas-based lesion disconnectomics in multiple sclerosis: a retrospective multi-centric study"

### Supplementary Materials

#### A. Extraction of individual tractograms

Image reconstruction and tracking strategies were chosen after what was described in (Yeh et al., 2018).

Reconstruction was performed using the Q-space diffeomorphic strategy (Yeh and Tseng, 2011) in MNI space with a mean diffusion distance ratio of 1 using DSI Studio (http://dsi-studio.labsolver.org). According to the methodology described in (Yeh et al., 2018), the whole-brain tractogram was built by first tracking 100’000 fibres at five different angular thresholds (40, 50, 60, 70 and 80 degrees) with a white matter mask defined by an anisotropy threshold of 0.6. Then, to allow tracking in the lowest portion of the brainstem, which would normally be outside the white matter mask, a region including the spinal cord in the lowest slice of the MR volume was manually drawn and 10’000 fibres terminating in this section were tracked for each of the five angular thresholds. Always following the tracking pipeline proposed in (Yeh et al., 2018), streamlines following the same path with a distance smaller than 1 mm were automatically removed.

#### B. Threshold analysis

In order to define the optimal binarization threshold for disconnectome graphs, the connection-level rank-rank correlation was computed for different threshold. Further, the Spearman correlation between large-scale features extracted from atlas-based and individual tractography based disconnectomes was also computed for different thresholds.

Supplementary Figure 1 reports the results of the threshold analysis. When looking at connection-level comparison in terms rank-rank Spearman correlation, a high agreement is observed between individual and atlas-based disconnectomes for both automated and manual lesion segmentations. This is especially true when lobe-wise connections are considered (Supplementary Figure 1A), whilst the correlation decreases for larger thresholds when considering individual connections (Supplementary Figure 1B). When considering the Spearman correlation between large-scale topological features (Supplementary Figure 1C), the agreement between the two methods globally improves for increasing thresholds but drops after 0.7. Topological features extracted from LeManPV lesion segmentation are following a similar trend compared to manual segmentation, despite from the transitivity which shows overall poor correlation when using LeManPV lesion segmentation.

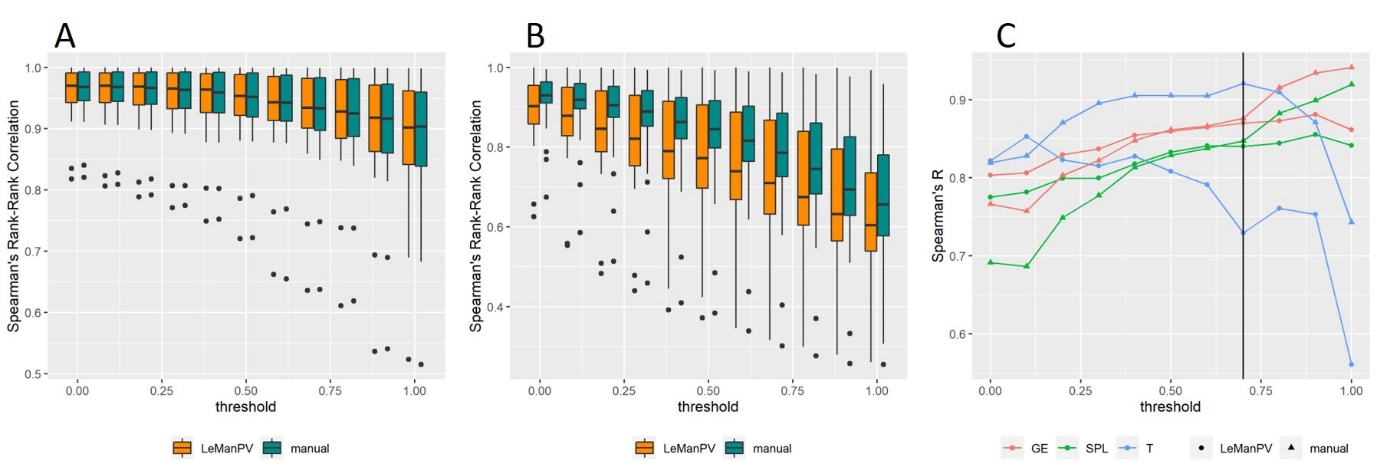

Supplementary Figure 1 Threshold analysis for graph binarization. A. Spearman correlation between ranks of lobe-wise connections from the atlas-based disconnectomes and individual disconnectomes for manual (green) and automated (orange) lesion segmentation. B. Spearman correlation between ranks of connections from the atlas-based disconnectomes and individual disconnectomes for manual (green) and automated (orange) lesion segmentation. C. Spearman correlation between large scale topological features extracted from atlas-based disconnectomes and individual disconnectomes for manual (triangle) and automated (dot) lesion segmentation. GE: global efficiency, SPL: shortest path length, T: transitivity.

#### C. Figures and Tables

| **Dataset** | **N_i_** | **N_0_** | **N_1_** | **N_2_** | **N_3_** | **N_4_** | **N_final_** |
| --- | --- | --- | --- | --- | --- | --- | --- |
| **Diffusion Cohort (patients)** | 43 | 0 | 0 | 0 | 2 | 0 | 41 |
| **1.5T cohort** | 220 | 0 | 7 | 1 | 2 | 2 | 208 |
| **3T cohort** | 582 | 0 | 5 | 0 | 4 | 77 | 496 |

Supplementary Table 1. Breakdown of patients in our multi-site framework after applying our applicability criterion as shown in Figure 3. N_i_ is the initial number of patients. N_0_ is the number of patients discarded due to poor registration quality. N_1_ is the number of patients discarded due to bad segmentation quality. N_2_, N_3_ and N_4_ are the numbers of patients with brain atrophy younger than 22 years old, between 22 and 36 years old and older than 36 years old respectively. N_final_, is the total number of patients eligible to compute atlas-based disconnectomes.

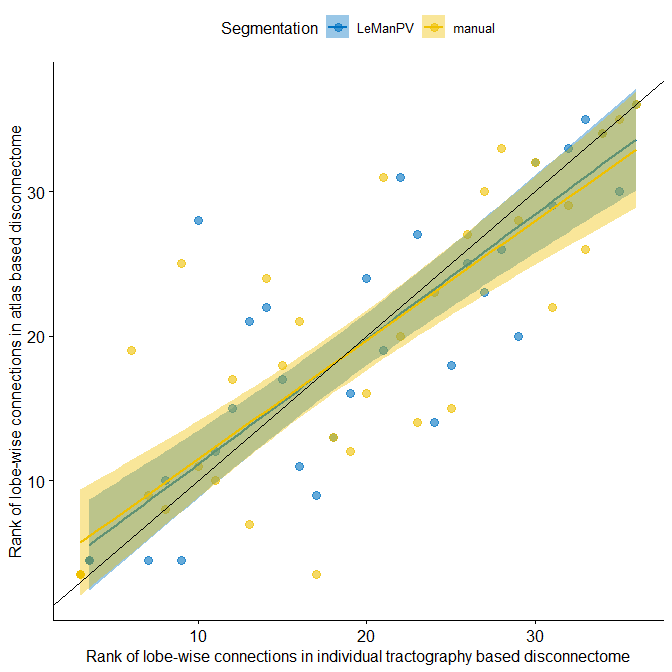

Supplementary Figure 2 Rank of lobe-wise connections from the individual tractography based disconnectome (x-axis) and atlas-based disconnectomes (y-axis) for both automated lesion segmentation with LeMan-PV (in blue) and manual segmentation (in yellow) for one patient of the diffusion cohort. Solid lines and shaded regions represent the regression line and 95% confidence interval.

Supplementary Table 2 Spearman correlations and respective p-values computed between relative absolute differences in topological features derived from atlas-based vs. DTI based disconnectomes, and age, brain volume and segmentation quality.

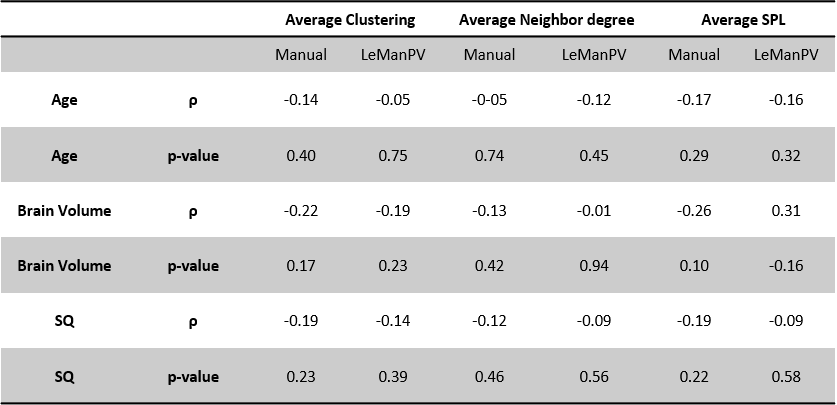

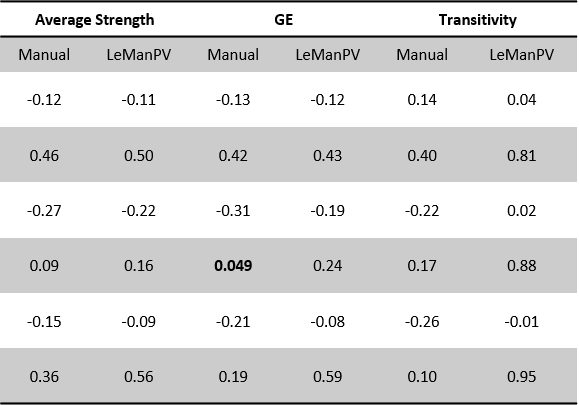

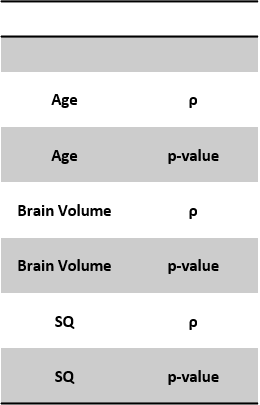

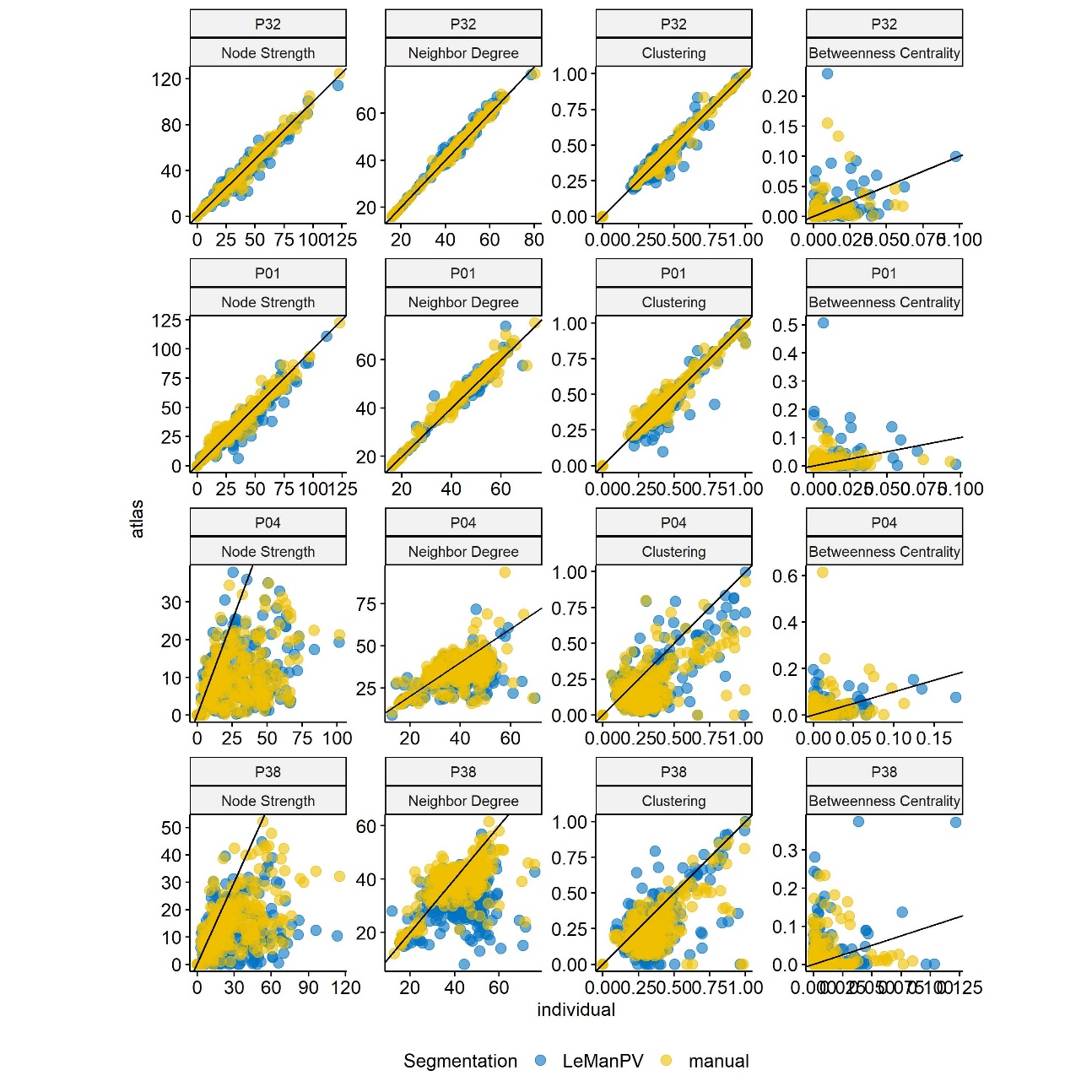

Supplementary Figure 3 Variation of small-scale topological features extracted from atlas-based disconnectome graph (y-axis) plotted against equivalent metrics extracted from individual tractography-based disconnectome (x-axis) for two patients showing a good agreement between the two techniques (P32 and P01) and two patients showing more differences (P04 and P38). The diagonal black line represents the identity function.
